# Disentangling the genetic causes of autism

**DOI:** 10.1101/2025.07.30.25332409

**Authors:** María M. Abad-Grau∗

## Abstract

**Background:** By suspecting that the scarce disease association results obtained by several genome-wide association studies, even in highly heritable diseases, could be due to wrong assumptions inferred from reference genomes but not true in affected individuals, we re-analyzed data from the Autism Genome Project consortium starting from the genotype calling phase.

**Methods:** We identified patterns of raw genotype intensities associated to the disease, mainly lack of hybridization. In light of it, we re-designed the polygenic models of individual risk and were able to explain most of the variance – AUC above 0.9 – in two independent datasets of whole exome sequencing (WES) provided by the Autism Sequencing and Human Autism Genetics consortia, by measuring differences in missing rates and Mendelian inconsistencies.

**Findings:** Analytical results when using a missingness test, pointed out to several highly significant positions, most of them having the autistic individuals as the group with the unknown variants. Results from Mendelian tests under our polygenic models, showed high accuracy and robustness as well, revealing a surprising pattern of higher rates of Mendelian inconsistencies –when considering rare variants– in unaffected offspring than in affected ones, even with affected and unaffected children belonging to the same family, and confirm autism as highly heritable. Missingness tests performed on copy number variation (cnv)s in sex chromosomes within array-based datasets, revealed that, in males, a missing cnv only in one gene (IL3RA) within pseudoautosomal region (PAR) 1 was able to explain the whole variance (*AUC* = 1) while in females it was also necessary but not enough another missing cnv, in gene IL9R, within PAR 2. Another cnv deletion, within a gene in chromosome Y very close to IL3RA, PRKY, is also highly associated to autism, and explains the large rates of aneuploidies in sex chromosomes of autistic individuals. Expression heatmaps performed with genes within autosomes found associated to autism, reveal cerebral cortex and testis as the regions most targeted by these genes. We also analyzed sequenced genotypes of Asperger and PDD-NOS individuals and found out that they are closer to individuals with typical development (AUC around 0.7) than to strict autistic ones (AUC above 0.9).

**Interpretations:** Although the origin of autism may be associated to deletions of a cnv at IL3R in males (common macrocephaly in autism may be explained by this), the strong linkage disequilibrium with PRKY, the close interaction between autosomes and sex chromosomes through evolution and large epistatic effects, result in several associated genes in autosomes strongly expressed in cerebral cortex and testis. In light of these results, autism may be confirmed as a spectrum of traits with thousand genome-wide genetic variants but with the origin in the deletion of a cnv in IL3RA in chromosome Y (PAR1), which may cause macrocephaly. Very often it is accompanied by the deletion of cnvs in PRKY, because of its strong linkage disequilibrium with IL3RA, which may cause, at least long-range, aneuplodies. The disease is transmitted to females because of PAR1 recombination but it also needs other genes in sex chromosomes (IL3RA together with IL9R in PAR2 are not enough) for the disease onset in females. Missingness tests showing almost no genetic differences between parents and affected offspring in autosomes (AUC around 0.5) seem to confirm that autism is a complex disease that requires not only the presence of the appropriate genetic variants, thousand of them perhaps, but environmental factors for the disease onset.

## Introduction

Since the end of the *Human Genome Project (HGP)*, more than 20 years of work of countless people and billion of dollars have been invested in *Genome Wide Association Studies (GWAS)* trying to find loci associated or in linkage with associated loci to complex diseases. Frequently, *de novo* variants are commonly presumed as possible explanations of results barely explaining a limited proportion of the variance, in studies that keep growing in sample size with the hope to increase the amount of significant loci, even in complex diseases with high prevalence and heritability.^1,2^ However, *Polygenic Risk Models (PRM)* from these diseases are expected to return high predictive rates of individual risk. Thus, the *Area Under the ROC Curve (AUC)* –a measure of accuracy in genetic risk predictors– should be larger than 0.9 in several complex diseases.^3,4,5,6^ In Autism, the disease we have focused on because of its high heritability (in fact, if an identical twin has autism, the likelihood that the other twin will also have is higher than 90%), maximum AUC should be between 0.997 –in Bangladesh, the world country with the lowest risk prevalence– and 0.999 –in Singapore, the world country with the largest prevalence–. No PRMs have been even dared to be used to predict individual risk of autism, as far as we know. In fact, even when using composite PRMs combining rare –*de novo* and inherited– with common variants, the variance explained barely reaches 4% .^7^

### Locus-based genotyping

Classic per-loci approach in genotyping may yield to biased results towards controls, mainly when using a reference genome as cases may have different genotypes related to the disease .^8^ Moreover, a considerable amount of loci do not show the common three-clusters pattern (AA/AB/BB genotypes) or they are very close to each other to perform a correct classification (see as an example figure 1a). The whole *Single Nucleotide Polymorphism (snp)* could be disregarded for not showing a clear three-genotypes pattern. In case the snp is kept, genotype call may be biased by the dataset. Figure a shows two extreme points in color: the genotype with the lowest intensity for allele B in green and the genotype with the larger intensity for allele A in red. Within the conventional per-loci approach, the whole snp could be classified as not a real snp and all the dots considered as BB genotypes. Therefore, the genotypes whose intensities are coloured could be considered as missing or as BB genotypes.

**Figure 1:**
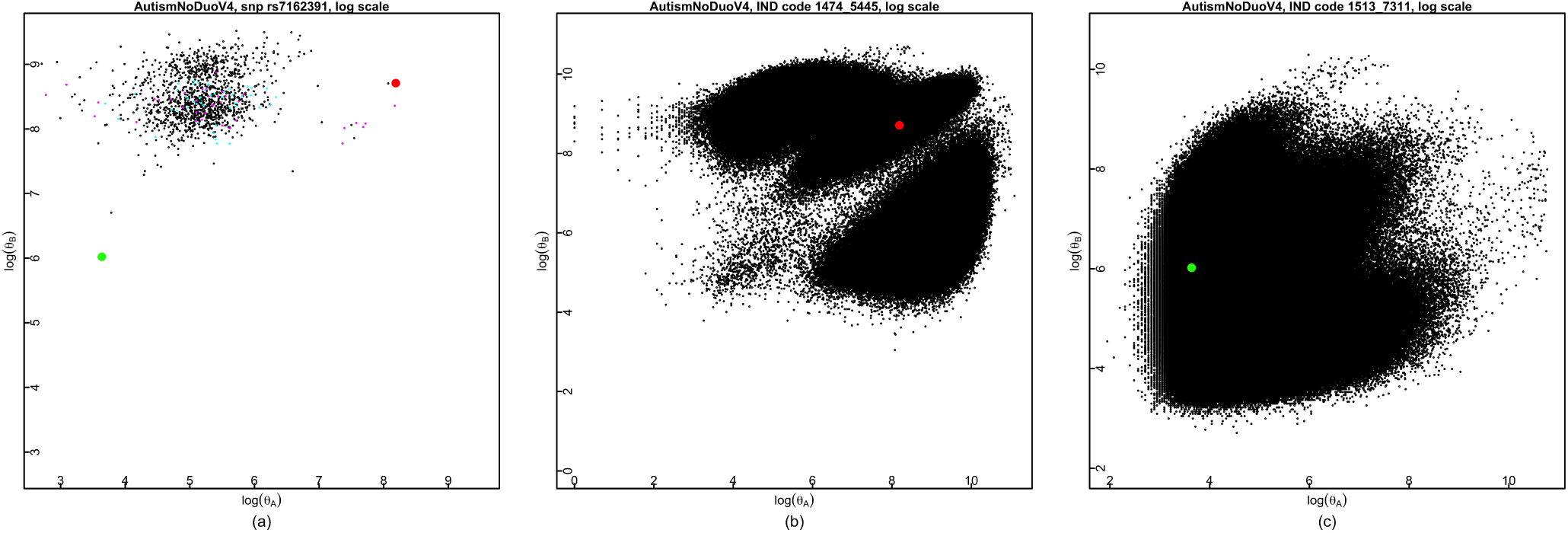
Scatter plots of raw intensities for the two alleles, *θ_A_, θ_B_* , one per axis, in log scale. Figure (a): Plot used in the conventional locus-based approach to call genotypes, in which all dots belong to a given locus (snp rs7162391 in the figure). It has been coloured in red the point with highest *θ_A_* and in green the one with the lowest *θ_B_* intensity. Figure (b): Plot used in an individual-based approach to call genotypes, in which all dots belong to a given individual. It corresponds to the individual with log intensities for snp rs7162391 – Figure (a)– shown in red. Figure (c): Another plot with individual intensities. It corresponds to the individual with log intensities for snp rs7162391 – Figure (a)– shown in green.

### Limitations of common Polygenic Risk Models

In an attempt of building individual risk predictors of complex diseases, taking advantage of GWAS data, different PRMs have been used. Most of them apply logistic regression on a *Polygenic Risk Score (PRS)*, a measure based on computing accumulated risk through different loci .^9^ A frequent way to reduce computational time when computing the score, is by selecting loci or by assuming independent contribution of loci or both. The first approach may disregard rare variants, the second one may bias (upward) effect sizes in presence of linkage disequilibrium (LD). To avoid this and other issues in the second approach, is a common procedure to select loci by using a reference genome to estimate LD, overall heritability, or allele weights. There are several problems to reproduce results with this approach though: from gross errors due to differences in allele coding that may be carried out from the genotype call step, to problems derived from population stratification when using datasets belonging to different ethnic groups.

A PRM based on machine learning may have the same gross errors, but there are many approaches to improve computational efficacy and generalization capacity (results reproducibility) without the need, and the consequent risk of increased bias, of using external information. They also can avoid overfitting by being parsimonious. Some exam-ples are näıve Bayes (which is actually equivalent to a low-biased PRS able to compute weights without using external information^10^), among Bayesian networks, or J48 among decision trees. The main advantage of J48, and decision trees in general, is to be white box models, i.e. the model learned can be easily understood by humans. Still all of these models are far away to achieve the expected AUC that they should reach when considering disease heritability and risk prevalence in many complex diseases, much less they will generalize in a validation dataset from, perhaps, a different population.

### Research in context

#### Evidence before this study

Autism is a polygenetic complex disease but there are many open questions: its etiology (it is only explained in less than 0.4% of individuals); their diversity and their association with thousand genetic variants; its difference in gender prevalence; its relationship with sex chromosomes and sex chromosome aneuploidies; differences between ASD, mainly between strict autism and Asperger or PDD-NOS

#### Added value of this study

Under the hypothesis that the lack of success in finding the genetic basis of most autistic cases could be due to difficulties to genotype new genetic variants, we have defined risk models by considering missing versus genotyped genetic variants instead of comparing different genetic variants among them and were able to correctly classify almost all cases and controls (AUC larger than 0.9 in four replication datasets) although it required many variants within all autosomes. We have also used trios (affected offspring and their parents) and quarts (a trio plus unaffected sibling) and found Mendelian inconsistencies more often between parents and unaffected than between parents and affected offspring. We found sex chromosomes (long-range) aneuploidies or at least chromosome-wide missing variants due to undetected copy number variations yielding to sex chromosome short-range aneuploidies were common in autistic offspring and their parents. Instead of hundred variants required to correctly predict autism from autosomes, we found that only one cnv in gene IL3RA in PAR1 was able to explain the whole variance (*AUC* = 1) in males, a gene influencing brain development and which may explain macrocephaly, a known indicator of autism. We also found a very strong association (AUC larger than 0.9) in a gene at chromosome Y very close to PAR1, PRKY, a gene associated with sex chromosome aneuploidies affecting gender genotypes (males XX and females XY) that may explain the large amount of aneuplodies, or chromosome-wide short-range aneuplodies, we found. In females we found that at least another gene, IL9R within PAR2, together with IL3RA, was required to predict autism and even with it, AUC did not reach 0.8 in one of the two dataset analyzed. The fact that only one gene is required to predict autism in males, and that females require other genes and PAR2 may probe why autism is not only a disease affecting males but, at the same time, it has a larger prevalence in males. In light of these results autism is confirmed as a polygenetic spectrum disease, with only one gene as a common etiology and many other in sex chromosomes in linkage disequilibrium with IL9R adding different phenotypic effects. We found genes associated to autism in autosomes are mostly expressed in cerebral cortex and testis, which may be explained because of the close interaction between sex chromosomes and autosomes. By building risk models able to differentiate strict than mild autism, we have also found that mild autism, such as Asperger or PDD-NOS is genetically closer to typical development than to strict autism. By realizing that our risk models were not able to differentiate autistic offspring from their same-gender parent, we confirmed that autism is not only a polygenetic spectrum but also a complex disease, i.e., genetic causes are necessary for the disease onset, but only with the right environmental factor the disease will develop.

#### Implications of all the available evidence

Individual predictors of autism may become soon available and ethical questions should be seriously considered. It is also important to understand that autism is a very wide spectrum disease; that most autistic cases may have a happy life and that, even strict cases, should be considered within the most absolute respect for a life that does not belongs to us, but comes to ours asking for protection, challenging and disarming ourselves and, perhaps, making ourselves better human beings.

## Methods

After realizing how far are we to build GRSs with these AUC values, we had the intuition that the clue could arise by questioning the first preprocessing steps, mainly genotype calling and *quality control (QC)*. We identified two procedures that could be better handled. One was the treatment of missing genotypes and *Mendelian inconsistencies (MI)*, that usually are not taken into account except for removing loci or individuals not passing QC, with the risk of removing a true effect. The other was related to the common per-loci approach used in genotyping methods, mainly in array-based GWAS. By using this approach and, even worst, computing prior distributions based on reference genomes, a high bias towards control genotypes is usually introduced .^8^ We defined new PRMs using missing and MI patterns in order to improve predictive accuracy. We also mixed the case/control approach –prone to type I error due to population stratification– with the family trios approach –designed mainly for *Transmission-Disequilibrium Tests (TDT)*, which turned out to have little power in complex diseases^11^– as a way to overcome the main issues of both approaches.

We have used genotypes from four different data sources, two using genotyping arrays (AGP-Human1M and AGP-Human1MDuo) and the other two (ASC and HAG) using *Whole Exome Sequencing (WES)*. We also have used data from IHMP as control genotypes for AGP sources. A summary of the datasets and data subsets used is shown in Table 1 (see Table 4 for a list of acronyms).

**Table 1:**
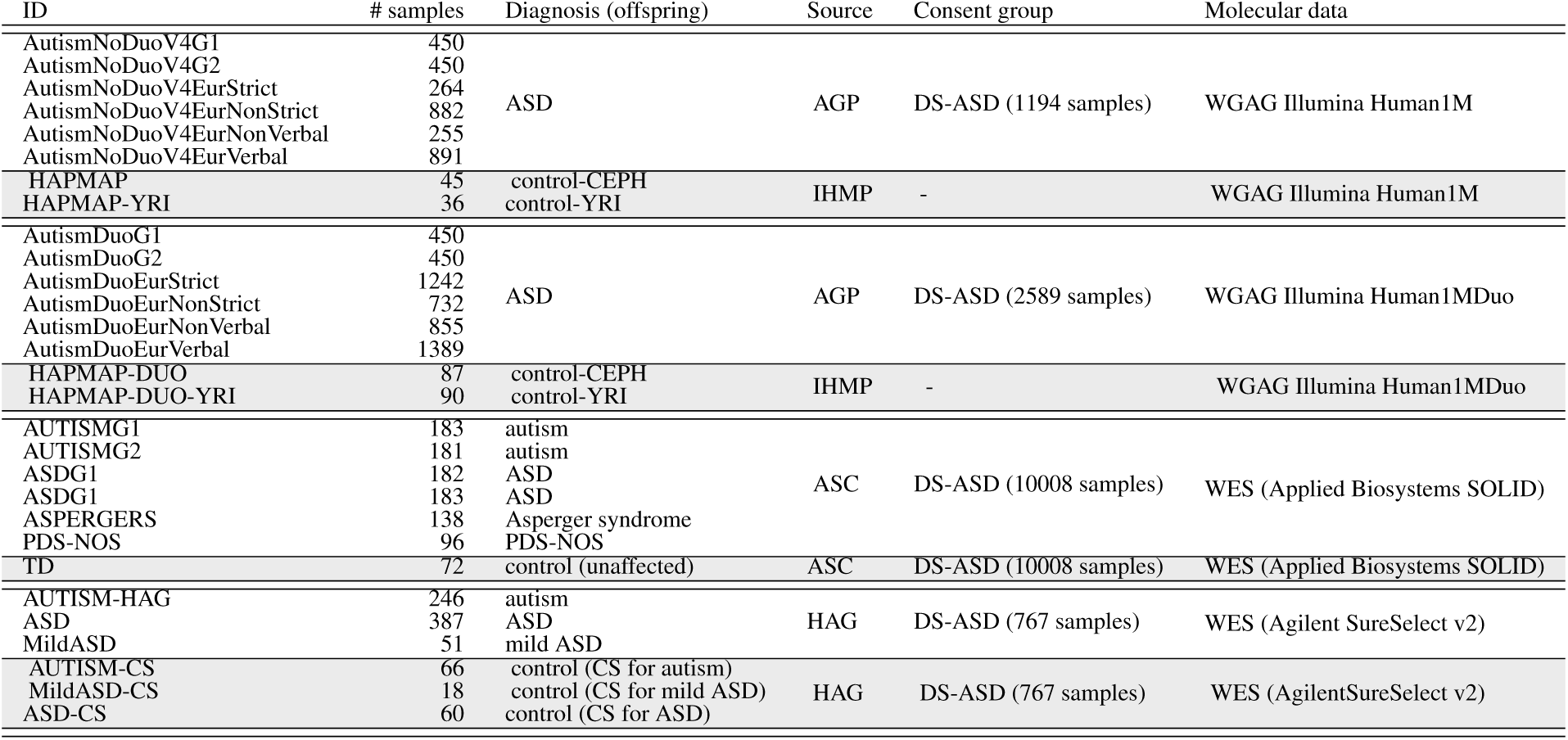
Datasets used in this study. Four groups of datasets, from different studies and molecular technologies have been used. For each group, controls are coloured in light gray. The first two groups are data from Illumina arrays and we used IHMP as quasi-controls, genotyped with the same Illumina array. For the other two groups, we used WES data from two GWAS^12,13^ were cases and controls were genotyped using the same procedure.

**Table 2:** Amount of loci screened by the four molecular designs used through all datasets (sex chromosomes included *Pseudo Autosomal Regions (PAR)* inside brackets) in this study and shared positions between the technology with the largest coverage (WES-SOLID) and the others.

| Genome-wide DNA technology | # Loci (chr.X — chr.Y — PAR) | # WES-SOLID Shared positions |
| --- | --- | --- |
| WES-SOLID | 3541420 (83366 — 1706 — 0) | - |
| WES-SureSelect v2 | 497016 (13432 — 270 — 0) | 323514 |
| WGAG Human1M (snps) | 812902 (29825 — 700 — 95) | 39827 |
| WGAG Human1M (cnvs) | 259914 (10272 — 1581 — 591) | 13101 |
| WGAG Human1MDuo (snps) | 799885 (26781 — 1204 — 187) | 37875 |
| WGAG Human1MDuo (cnvs) | 399302 (18810 — 3433 — 792) | 19684 |

### Treatment of missing genotypes and Mendelian inconsistencies

We found genotype calling methods usually removed true associations by building prior models based on common healthy populations, mainly using data from the *International Hapmap Project (IHMP)*^14^ or the *1000 Genomes Project*.^15^ By trying to study the pattern of missing and Mendelian inconsistent (MI) loci in different complex diseases, we found that these patterns were far a way of being at random. Therefore, they should be considered as part of the genetic risk models, instead of being disregarded by too much stringent QC procedures. Regarding missing data, we found out that, when using a case dataset to build prior models in a genotype calling method instead of a control or reference data set, the proportion of missing calls was larger in a control dataset.^8^

There are at least two different sources of missingness, that should be differentiated: one is an uncertain call when raw intensities lies in between two of the three classic genotypes (homozygous wild type, homozygous mutant and heterozygous) and the other is an uncertain call because raw intensities for the two alleles are too low to sustain the underlying hypothesis of an individual having one of the three possible genotypes (non-hybridized probes). Genotype call algorithms usually do not make this difference and most *Genome-Wide Asociation Studies (GWAS)* QC procedures disregard whole individuals or loci when there are two many values not compatible with one of the three genotypes identified by the calling algorithm. When upgrading to *Next Generation Sequencing (NGS)* technologies, at the end of a highly greedy process in terms of computer and human resources, we always lose rare variants and consider then as missing genotypes, although the amount of losses depends on a trade-off between cost saving and effectiveness. Again several loci or individuals are disregarded if there are missing genotypes, and analyses performed on the genotypes keep either disregarding missing genotypes or performing imputation just to avoid algorithm halting. In our honest opinion, the question, not well-handled so far, is whether the missing values are really informative and associated to the trait analyzed.

The second source of information that can be erroneously disregarded, is the one related to Mendelian inconsistencies. A common test used in QC procedures, mainly when using family trios (an affected offspring and their parents), is a Mendelian test, with the intention of disregarding loci or individuals with high levels of Mendelian inconsistencies as it may reveal false paternity. The problem also arises with the way to handle those sparse inconsistent genotypes, mainly annotated as missing. Or, even worse, we may not correctly ascertain whether large amounts of Mendelian inconsistencies really means false paternity or are a true signal of highly structural mutations.

### Individual-based genotyping

By using an individual-based approach, genotype call is performed for a whole individual at a time. Hence, plots are built with all loci for each individual (see the plots of two individuals in figures 1b and 1c). This way, the genotype in red would clearly be ascertained as an AB genotype (figure 1b) and the one in green as a BB genotype (figure 1c). Moreover, with this individual-based approach, important differences between different individuals arise, and help to understand that quantile normalization procedures commonly used to reduce technical variations between samples, may barely improve what seems to be a too much reductionist locus-based approach.

As an example, the two individuals in figure 1b and figure 1c have a very different shape, perhaps because DNA replication went faster within the array bead corresponding to the sample from individual shown in figure 1b than within the one corresponding to the sample from the individual shown in Figure 1c (mean intensities are larger in figure 1b).

### Genotype calling

For our goal of detecting non-hybridized snps that can point out to an alternative variant, it would be more important to have low bias towards the reference genome or towards the majority of the dataset being analyzed than low error rates. Therefore, we developed what we have named *Individual Genotype Call (IGC)*, a genotype call procedure able to identify as missing only genotypes compatible with non-hybridized probes. The algorithm works under a per-individual basis avoiding both, normalization ^16^ and early removal of variants or samples whose anomalies may be disease-associated. We were not interested in simplistic statistical summarization, neither in finding common patterns that may remove outliers, but in a wide and deep search of low-effect variants, mainly in the case of spectrum disorders with a high and diverse amount of genetic variants affecting different individuals, as in ASD. Therefore we did not utilize neither a reference sample or the remaining individuals under the current analysis. Another advantage of performing genotype calling for one individual at a time would be the reproducibility of results in completely different samples and the feasibility of a direct development of a disease profiling test just requiring the genotypes of the offspring or their parents, ascertained one by one. Due to the very different log-raw-intensity scatter plots (we will refer to them as gentprints) between different individuals, we did not succeed in the design of an accurate and robust clustering method, either by using statistical distributions or computing vision methods for object recognition. As an example of a computer vision method, we tried to perform genotyping from a 3D gentprint using frequencies as the *z* axis and a Hessian matrix (second derivative) to propagate from cluster peaks to valleys (see in figure 2a a 3D gentprint map for a case where most genotypes are easily recognized by view as belonging to AA, AB or BB group). The inclusion of a missing group made clustering more difficult to succeed. In fact some genotypes were wrongly classified as missing (figure 2b). In order to understand which points in a gentprint should be classified as missing due to both intensities being too low, we made coloured gentprints and used a different color for autosomes (black), chromosomes X (orange), Y (cyan) and PAR-XY (magenta). As females do not have Y chromosome, cyan dots in female gentprints can give an idea about the position where non-hybridized points in any other chromosome should be placed within the gentprint (see figure S4 for a trio from AGP and figure S5 for a trio from CEPH IHMP, both from array Illumina 1MDuo). Gentprints in these figures also give an idea of the large plot variability within the same family and support the idea of using an individual-based approach instead of the position-based conventional approach with a normalization procedure.

**Figure 2:**
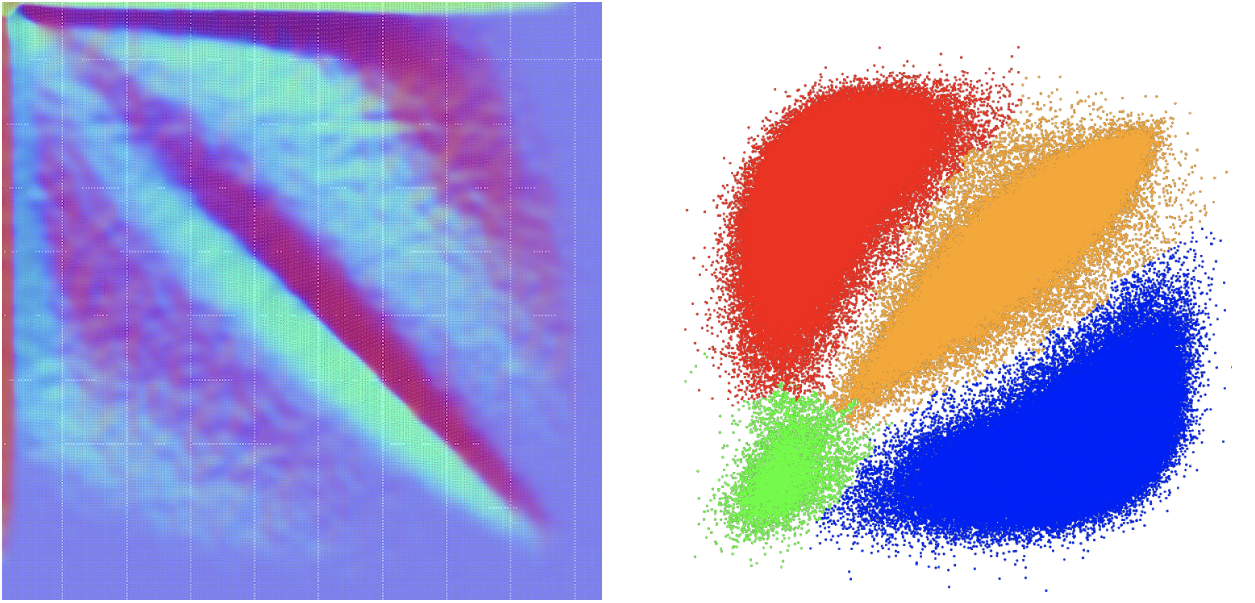
GentPrints to show a computing vision method for genotype clustering. (a) A 3D gentprint map (Y axis is inverted because it is a graphical image, not a statistical plot) where it can be observed the intuition about the genotype calling method. (b) A gentprint with already classified genotypes as blue (AA), orange (AB), red (BB) and green (missing). Some genotypes classified as missing seem to belong to known alleles.

As an alternative, we decided to use an artificial vision approach to build a model. A human expert did the first phase of the model training, by performing genotype clustering by view of the gentprints of 40 samples coming from all the arrays from which we will later perform genotype calls (we used Illumina Human 1M and Human 1M Duo for this study). Figure 3a shows an example of an “irregular” gentprint, with off-center AB cluster. In a second phase during model training, the model was applied on other 180 samples and the human expert just accepted or corrected the clustering of each new image (it was a yolov8n model trained with Roboflow instance segmentation with 96.1% sensitivity and 96.1% precision in the test data subset). The model was applied on new gentprints in order to have the four irregular polygons (an example of model inference can be seen in figure 3b). The computer vision watershed algorithm is utilized afterwards, considering the points inside polygons as local minima, in order to classify all the genotypes (see figure 3c). It has to be observed that samples used to build the model do not bias genotyping outcomes the way they do when using the conventional by-locus approach. In fact, by using an instance-based approach instead of a per-locus approach, the model focuses on each individual and is much less influenced by other individuals, helping to identify four regions by adapting itself to very different maps. It just learns to identify always 4 labels, even if there are very few points in some of the clusters (usually the missing genotype) and it does not classify as missing those points with ambiguous genotypes (points between AA/AB or between BB/AB clusters). Instead of that, it uses the Euclidean distance to initial clusters for genotype call. Therefore, we can now be more confident in that missing genotypes are mainly due to lack of hybridization, which is consistent to an individual having completely different variants at a given locus than the reference alleles considered by the array designers.

**Figure 3:**
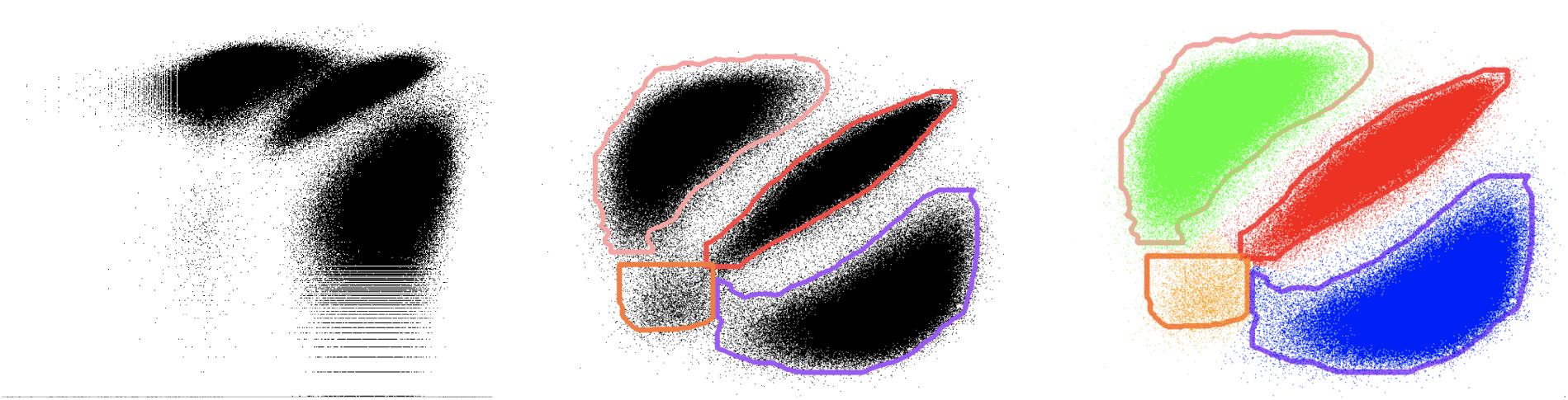
A summary of the IGC procedure to perform genotype calling on a given individual. First we produce an individual log-raw intensities map for all positions and chromosomes (a). Then we infer clusters using yolov smart polygons (b). And finally we classify the points outside de polygons as well by using the watershed algorithm.

### Technical bias during genotyping

We observed, regarding array-based data, a large difference in the amount of non-hybridized loci between different array configurations, differences that may also exist when genotyping is performed from different labs, even using the same array model. Non-hybridized loci is the source of missingness that points out to a *Hidden Hidden (HH)* genotype, i.e., a genotype not being neither AA, BB or AB but having two different variants, two copies of a different variant or deletion of the two alleles. In fact, there is a much larger amount of HH genotypes in Human1M-Duo than in Human1M. These differences mainly apply to autistic offspring and their parents. Thus, when using IHMP data from CEU and YRI populations as control data, both from the same Illumina arrays, this pattern was hidden as there were higher amounts of intensities eliminated by the scanning systems in HapMap raw intensity (idat) files than in AGP datasets. This is a very important result, because it shows the need to use the same array in cases and controls for comparative purposes. Moreover, still using the same array, the first steps to return intensities may bias final results if cases and controls are processed at different labs. Therefore, when using a case/control approach, it becomes mandatory to use exactly the same procedure in the analytic platform to extract bead intensities from the fluorescent image produce by the labeled DNA. Differences in procedures yield to technical-based differences in missingness rates that may produce spurious association.

For WES-based genotypes, we used processed data returned by the last step of the two studies that produced them .^12,13^ This means the sequenced data we used already passed a QC process. Both of them used Picard and BWA for alignment and reads mapping, and GATK for SNP/variant calling (details of specific procedures are provided in their respective supplementary material). We are aware of the problems of using the data already processed. Mainly, missing genotypes depend on the reference genomes used. However, for this study, as they used exactly the same procedures for cases and controls, is enough to have these data in order to test whether the unknown variants, annotated as missing genotypes, may be associated to the disease or not.

### Family trios vs case/control approaches in complex diseases

Although family trios require greater efforts to be collected, they seem to be preferred to classic control/cases approaches in GWAS of several complex diseases, mainly because they get rid of type I errors derived from stratified populations. However, perhaps we should challenge the way we use family trios once we consider missing data likely point out to hidden rare variants. So far, because of the lack of positive results with specific association tests for family trios, such as the TDT, the main solutions have been to increase sample sizes and to look for *de novo variants (DNVs)*. Still, neither of these solutions seem to be able to explain the majority of the variance in several complex diseases. We should not forget that a complex disease is still an inherited disease, one which requires some environmental factor for the disease to onset – with a very high heritability rate in the case of autism– and DNVs should not be that important for a highly heritable disease.

### Missingness-based and MI-based Polygenic Risk Models

We defined the simplest missing-PRM we could think about without any prior distribution: an accumulative model from binary values at each locus, with no weights. We could use a PRS (logistic regression) or a PRM able to handle numerical attributes, such as decision trees or decision forests, neural networks, support vector machines, etc. There is only one input variable per chromosome, representing the amount of missing data an individual has at a given chromosome, or only one input variable at all if all chromosomes are collapsed. We actually built a different classifier by chromosome and one classifier with the total amount of missing rates among all the chromosomes, as the unique input variable. As it is a very simple supervised classification problem, we opted for a J48 decision tree, in order to use a white-box model. The models will be just a threshold or cutting value for the input variable and the class for each half of the input variable.

In order to discover whether the amount of missing genotypes influences MI and disease association and the impact of inheritance and the novo missing variants in autism, we defined some variants of missing and MI-PRMs (see table 3 in Methods for a description of all the PRMs used). We assume missingness may be due to either rare (unknown) variants, cnvs, deletions, inversions or other structural variants. We also defined a PRM with heterozygosity proportions of loci per individual, even if this PRM is prone to type I errors in presence of population stratification (see Methods).

**Table 3:** Description of the seven PRMs defined in this study. All the counts/proportions are computed either by chromosome or using all or a sub-group of chromosomes (v.g. autosomes). In order to use counts instead of proportions (in missing and MI PRMs), cases and controls need to be produced by exactly the same molecular technology.

| PRM acronym | PRM meaning | PRM description |
| --- | --- | --- |
| missing | Missing-based PRM | Rare variants measured as counts of missing loci for each individual |
| MI | MI-based PRM | Inheritance including rare variants measured by the proportion of Mendelian inconsistencies for each individual |
| hetero | hetero-based PRM | Heterozygosity measured as counts of heterozygous genotypes for each individual |
| inh-missing | inherited missing-based PRM | Inheritance rare variants measured as the proportion of missing offspring genotypes given at least one missing parent |
| deNovoMissingMI | *de novo* missing-based PRM | *de novo* variants measured as the proportion of MI due to missing offspring genotypes given both parents have known genotypes |
| repairingMI | Repairing MI-based PRM | DNA repairing rates measured as the proportion of MI due to known offspring genotypes given both parents have rare variants (missing genotypes) |
| basicMI | Basic MI-based PRM | MI in absence of rare variants measured as the proportion of MI given offspring and their parents have known genotypes |

An alternative genotype data that can be also used to build a PRM, which avoid gross errors as well due to differences in allele coding, especially when the control and case datasets come from different sources, is the number of heterozygous loci per individual. We referred as “hetero-PRM” to a PRM built with this information. For WES-based datasets, we considered all heterozygous genotypes, regardless the variants, some of which are not SNPs but polymorphic micro-satellite markers. Hetero-PRMs (assuming, in the array-based datasets, that the genotype call algorithm is good enough to classify as missing non-hybridized genotypes), can show some complementary information in order to identify loci associated to a given trait. However, this PRM is not specific at all, as it may as well return high AUCs in presence of population stratification, the most extreme situation being whenever case and control samples come from different populations. For this reason we included this PRM. We used it together with two highly stratified data subsets combined on purpose: the two array-based autistic offspring and their parents (AutismDuo and AutismNoDuoV4) as cases and, as they are mainly Caucasians, IHMP Yoruba offspring and their parents.

### Sex chromosomes

We used plink to impute gender from chromosome X (inbreeding coefficient) and chromosome Y (missing frequencies). We first chose threshold from the control data subset of each data source in order to have all control individual with the same imputed and pre-assigned gender (except outliers). Within AutismDuoG1 there was more than 50% of fathers and offspring with several loci compatible with karyotype 48,XXYY. This result is not confirmed in the second data subset within the same data source neither within the other data sources, still all datasets have larger rates of aneuploidies in cases than in controls. Therefore we believe results from AutismDuoG1 cannot be explained only as array design and lab procedure bias but to some unknown genetic structural variant of autism associated to sex chromosomes, which together may cause those results. Supplementary figure S4 shows a trio gentprint in which both parents have a karyotype compatible with 48,XXYY. This trio gentprint can be compared with a normal one, as the one shown in supplementary figure S5 from a HapMap family trio.

### List of acronyms

**Table 4:** List of acronyms.

| Acronym | Meaning |
| --- | --- |
| AC | Autism Carrier |
| AGP | Autism Genetic Project |
| AGP-DUO | AGP source datasets using WGAG Human1MDuo |
| AGP-NODUO | AGP source datasets using WGAG Human1M |
| ASC | Autism Sequencing Consortium |
| ASD | autism spectrum disease |
| CEPH | Centre d’Etude du Polymorphisme Humain |
| CNV/cnv | Copy Number Variation |
| CS | control siblings |
| DNV | *de novo* variant |
| DS-ASD | disease specific (autism spectrum disorder) |
| HAG | Human Autism Genetics |
| Human1M | Illumina genotype array Human 1M (1072820 oligos/SNPs) |
| Human1MDuo | Illumina genotype array Human 1M Duo (1199210 oligos/SNPs) |
| IGC | Individual Genotype Call |
| IHMP | The International Hapmap Project<sup>14</sup> |
| LRLD | Long-Range Linkage Disequilibrium |
| PAR | Pseudo Autosomal Regions |
| PDS-NOS | pervasive developmental disorder not otherwise specified |
| POT | parent-offspring trio |
| SNP/snp | Single Nucleotide Polymorphism |
| TD | typical development |
| WES | whole exome sequencing |
| WGAG | whole genome array genotyping |
| YRI | Yoruba (Ibadan) |

## Results

### 1. Results

We investigated whether autism can be predicted from genome data, mainly taking into account missing genotypes and MI, as they may point out to rare variants. For array-based datasets, we designed a genotype calling algorithm able to differentiate the two main sources of missing data: because of uncertainty between possible genotypes or because of a lack of hybridization (see Methods). We built genome-wide (exome-wide, for WES datasets) or chromosome-wide PRMs, for what we needed case/control datasets. However, to control upward-biased outcomes, due to population stratification, linkage disequilibrium, unbalanced case/control datasets or molecular procedures (see technical bias during genotyping, in Methods), between cases and controls, we implemented several methods, including the use of different types of PRMs built with the genomes of family trios. We have *parent-offspring trios (POT)*, with affected offspring for all the sources used. We also looked for POTs with unaffected offspring and their parents to use both, case/control and family trios approaches. One of the data sources used (HAG) has family quartets (quads), i.e., affected offspring, unaffected sibling and their common parents (an invaluable approach to avoid population stratification and, at the same time, to study whether a variant found in association with the disease –truly associated or in linkage disequilibrium with another variant– has been inherited from parents or is a DNV).

#### 1.1. Missingness-based and MI-based Polygenic Risk Models

When the conventional, allele-based, models have not succeeded, *missingness-based PRMs (missing-PRMs)* and *MI-based PRMs (MI-PRMs)* may be very important to identify variants, even if they are not known, related to any disease. Their utilization of missing/MI pattern information, disregarded by the common allele-based models, have become crucial to reach AUC rates close to their expected values. To build our PRMs we just counted the proportion of missing/MI per individual, either for each chromosome or for all chromosomes together (for MI-PRMs, we need parental genotypes as well). We computed AUC of J48 decision trees in a 5-fold *cross validation (cv)* approach using autosomes (we show average AUC between-chromosomes in most plots) in the autism training/test subset pair from each of the four data sources used in this work (AGP Human1M, AGP Human1MDuo, ASC-WES and HAG-WES, respectively). For arrays (AGP Human1M, AGP Human1MDuo), we also composed on purpose a highly population-stratified pair of data subsets, by using IHMP Yoruba as quasi-control data subsets (most cases are Caucasians) in order to check whether PRMs are upward-biased under population stratification (see table 1). Results, in figure 4, confirm that missing (red-coloured) and MI (blue-coloured) PRMs reach high AUCs in most data subsets close to what it should be expected. All the other PRMs (see a short description in table 3) were not expected to capture the majority of the variance, but to help to better understand (1) the data used, (2) results returned by MI and missing PRMs and (3) different ways to select loci to improve accuracy as it will be shown below. We compared these results with AUCs returned from PRMs built from less severe ASD cases, such as Asperger, *Pervasive Developmental Disorder Not Otherwise Specified (PDD-NOS)*, mild ASD or non-strict autism (in fact three of our four sources of data used different ways to classify ASD severity). Figure 5 shows results from mean AUCs among autosomes. They are quite different between data sources: data from arrays (left panel, AutismNoDuoV4/AutismDuo) are not able to differentiate autism according to strictness or verbality, HAG-WES (right panel) show also low AUCs, while ASC-WES show the best relationship with phenotypes (we still need to understand the reasons for these differences). Asperger and PDD-NOS individuals seem to be genetically close to each other with almost no differences in our data (AUC close to 0.55 in MI and missing PRMs). Moreover, both of them are closer to *Typical Development (TD)*, the group we used as controls for the ASC source (AUC close to 0.7 in both missing and MI PRMs), than to autistic cases, with a very high AUC (almost 0.9 in the MI PRM), which means they are highly differentiable based on their genomes. We also aimed to understand the extent of genetic inheritance versus DNV in autism PRMs. For this, we built PRMs to see whether they could differentiate case individuals from their closest relatives used as controls. Figure 6 shows results from mean AUCs among autosomes. A first result (figure 6, left panel) is that there are no differences when comparing genotypes from cases and their parents using hetero and missing PRMs (MI-PRM does not have sense under this approach). The result (AUC *≈* 0.5) is the same for all the sources used. This result is consistent with a complex disease, such as autism: some environmental factor may trigger the disease, given the individual have the (poly) genetic variants required. Therefore, parents carry and transmit autism-associated variants without being, most of them, affected. Another interesting result, in agreement with heritability in autism and other ASD, is that genetic differences between affected and unaffected offspring are much larger than between affected and their parents, with differences increasing with disease severity (figure 6, right panel: AUC close to 0.9 between autistic offspring and their unaffected siblings in MI-based PRM but also very large in missing, inherited missing and repairing missing).

**Figure 4:**
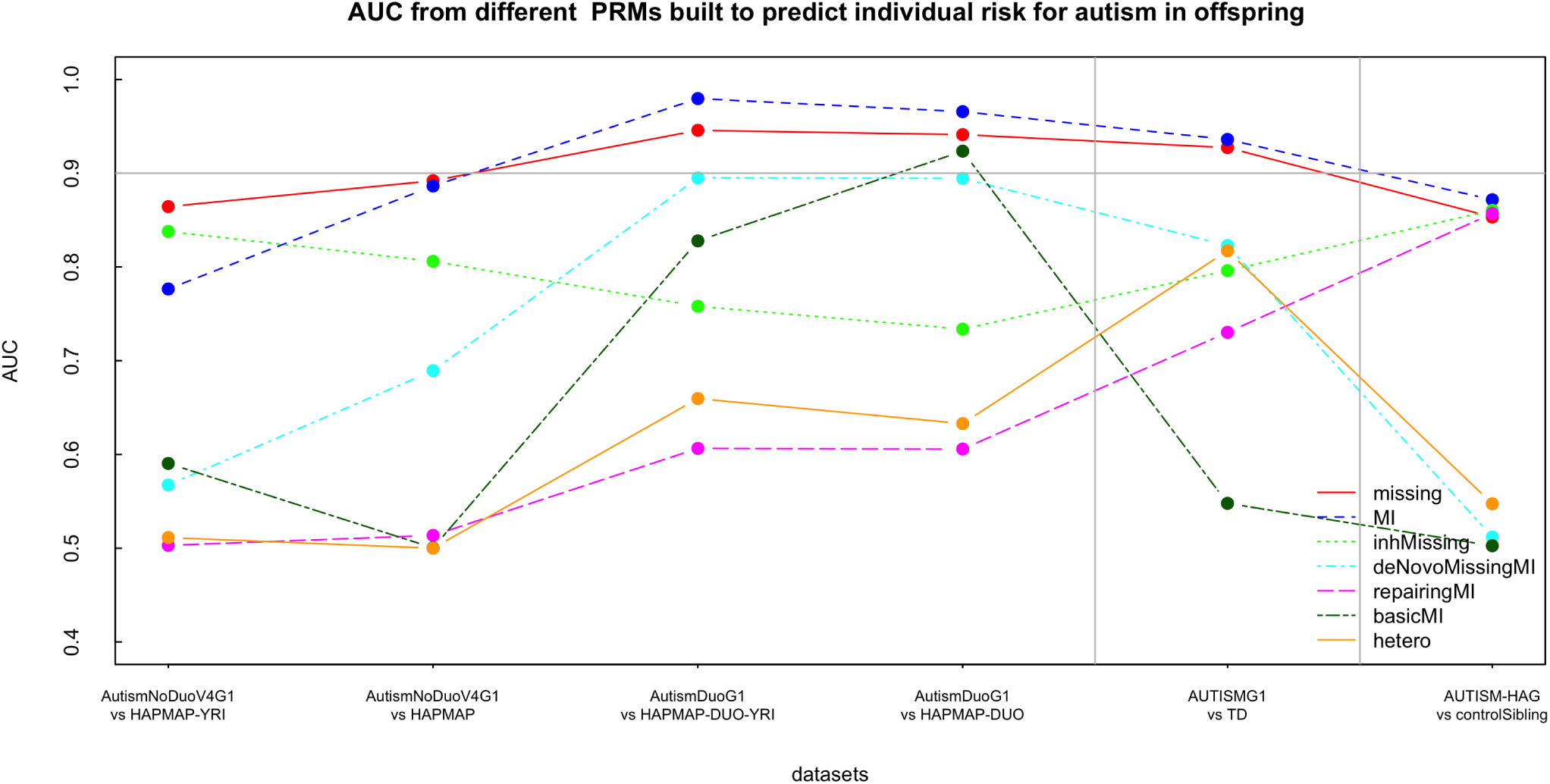
AUC results (Y axis) using the mean from 22 J48 decision trees (one per autosome) to predict offspring affectation risk. We built decision trees for missing (red), MI (blue), inh-missing (green), deNovoMissingMI (cyan), repairingMI (magenta), basicMI (dark green) and hetero (orange) PRMs using counts/proportions from offspring in the four data subsets of autistic offspring, unaffected controls and their parents (X axis).

**Figure 5:**
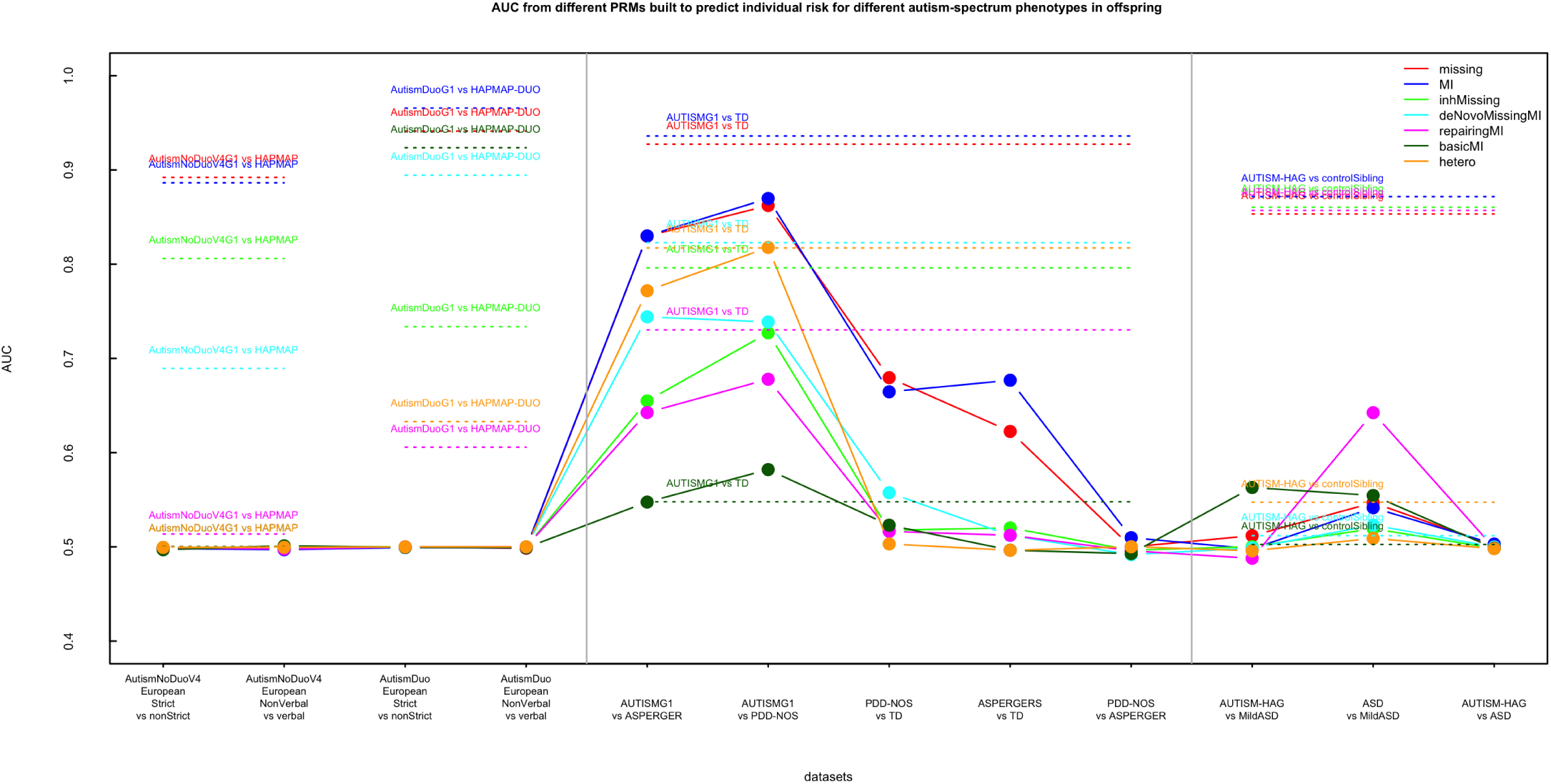
AUC results (Y axis) using the mean from 22 J48 decision trees (one per autosome) to predict offspring affectation risk. We built decision trees for missing (red), MI (blue), inh-missing (green), deNovoMissingMI (cyan), repairingMI (magenta), basicMI (dark green) and hetero (orange) PRMs using counts/proportions from offspring in different case/control data subsets (X axis) where cases have either autism or less-severe ASD, in the way phenotypes were collected by the four different sources whose data are being used in this work: strict/non strict and non verbal/verbal in Duo and NoDuo AGP, autism/PDD-NOS/Asperger in ASC and autism/ASD/mild ASD in HAG .

**Figure 6:**
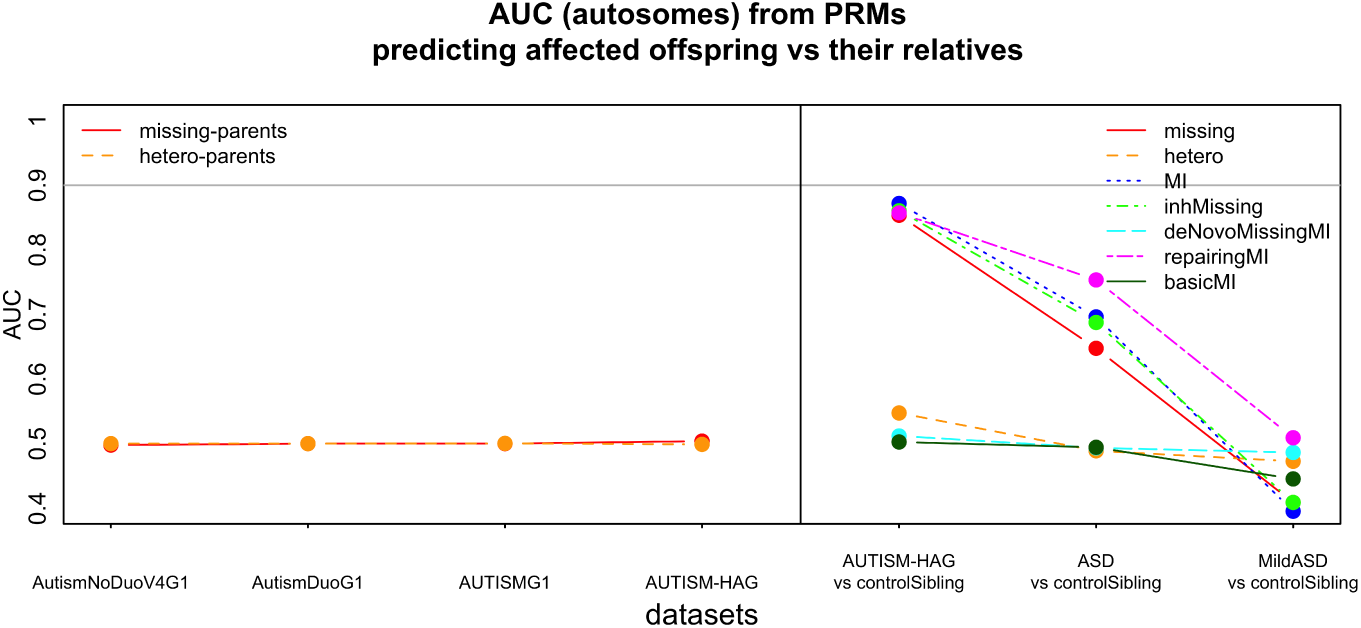
AUC results (Y axis) using the mean from 22 J48 decision trees (one per autosome) to predict offspring affectation risk. We built decision trees for missing-parents (red), hetero-parents (orange) PRMs (left side) and missing (red), MI (blue), inh-missing (green), deNovoMissingMI (cyan), repairingMI (magenta), basicMI (dark green) and hetero (orange) PRMs (right side) using counts/proportions from offspring versus parents (left side) or control siblings (right side). We used different case/control data subsets (X axis) with cases and their parents (left side). We used data from the WES HAG source to model counts differences between offspring and their unaffected siblings (right side).

#### 1.2. Filtering and replication

In order to replicate these results and to test whether AUC increases with loci filtering, we first tested whether differences in counts for each loci within all the PRMs built between affected and unaffected individuals were statistically significant (we used *χ*^2^, *χ*^2^ with Yates correction and a *N −* 1 two proportion tests). Table 5 contains results under Bonfferroni correction for the main affected-unaffected subsets within the four sources of data (HAG-WES, ASC-WES, AGP-Human1M and AGP-Human 1MDuo). Results show the number of significant loci when rates are larger in data subsets of affected individuals (Case columns) and when rates are larger in data subsets of unaffected individuals (Control columns). The best and highly significant results are given by the HAG-WES source (first set of three rows) but they are also mainly confirmed by ASC-WES dataset (second set of three rows): all (inherited missing PRM) and almost all (missing PRM) significant loci point out to more missing genotypes in affected individuals and all (hetero, *de novo* missing MI, MI and repairing-MI PRMs) point out to more hetero/MI in unaffected than in affected individuals. However, the number of significant loci is very different. In HAG-WES, the most informative PRMs are inherited-missing, MI and repairing-MI, with a similar number of significant loci (around 30000) –which are mostly shared between PRMs (data not shown)–, which yields to an immediate interpretation: there are rare variants in thousand loci in the genomes of autistic individuals (missing PRM), most of them inherited from their parents (inherited-missing PRM). Unaffected individuals do not share variants with their parents (MI PRM), they seem to have repaired the rare variants of their parents. Our conjecture is that parents have a source of structural variation, genome-wide duplicons, inversions or short-range aneuploidies which are classified as rare variants (missing) by the SNP/variant calling algorithm in data from WES. This conjecture is supported by the fact that there are not any statistically significant loci when using repairing-MI PRMs and very few when using inherited-missing PRMs, in array-based datasets (the third and the last set of three rows in the table). In fact, most algorithms used to call genotypes, included ours, would not differentiate between single, duplicated or inverted allele variants.

**Table 5:**
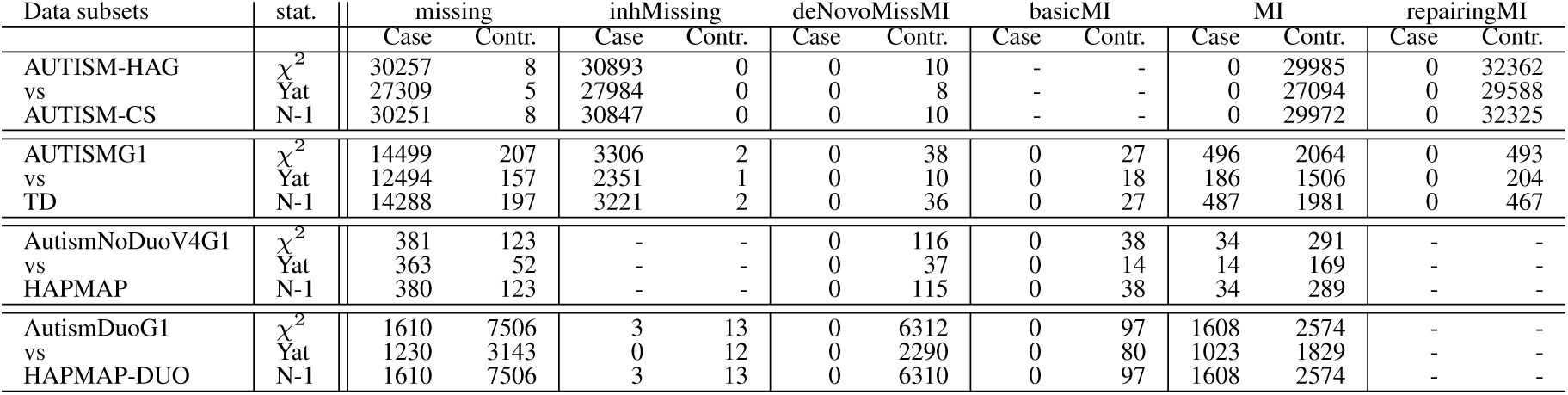
Counts of significant loci (p val*<* 0.05, Bonferroni correction) associated to autism (offspring) for the main training-test data subsets of each data source used at the training-test step for the 6 different PRMs (columns) and three different statistical tests to measure case/control differences: *χ*^2^, *χ*^2^ (Yat in the table) and N-1 two proportion (N-1 in the table) tests. Contr. (controls) means the number of significant loci under the alternative hypothesis of control (or, in general, the second data subset/trait) having higher missing/MI rates and Case (cases) means the number of significant loci under the alternative hypothesis of higher proportions of missing/MI in cases than in controls. In case of not any significant positions found at all, a hyphen is shown under both (Contr. and Case) columns. Results should be compared between tests but only within data subsets, as sample sizes are different between data subsets (see table 1) and loci counts are different between technologies (see table 2).

For filtering, we used the best results, AUTISM vs their control siblings (AUTISM-CS) from the HAG-WES source of data to filter loci (‘HAG-WES’ filter), due to their highly significant results. Therefore, loci from the other three sources of data were filtered according to these loci (we divided loci using 10 bins from their *log*(*p*), being *p* the *p* value). We computed AUC using all loci as well. For the HAG-WES dataset, due to lack of data, we used ASD vs their control siblings (ASD-CS) as training/test datasets and AUTISM vs their control siblings (AUTISM-CS) as replication subsets. We built the PRMs with filtering using the pair ‘AUTISMG1 vs TD’ (ASC-WES dataset) in order to use an independent filtering subset (‘ASC-WES’ filter). We also used the ASC-WES source as a second way to filter data from arrays. Figure 7) show results for the four dataset with p value selection using *α* = 0.05 with Bonferroni correction. All results were obtained by measuring AUC in the replication data subsets. X-axis shows AUC for each filter bin, and max AUC among autosomes without filtering (right-most value). AUC results using ASC-HAG for filtering are shown in green (missing-PRM) and dark green (MI-PRM). AUC results for filtering using ASC-WES are shown in red (missing-PRM) and blue (MI-PRM). All these results show very high AUCs, including results from ASC-HAG (plot b), for which we had quads to control any chance of population stratification. Regardless filtering, all these results show that genomes of autistic individuals can clearly be differentiated from healthy counterparts by just counting either missing genotypes (more missing in cases pointing out to rare variants or deletions) or MI (more MI in controls, pointing out to larger DNA repair in controls). The fact that results are almost always higher without filtering (the exception is in AGP-DUO) may point out to the absence of any loci within autosomes causing autism but only in linkage with putative loci, most likely within sex chromosomes.

**Figure 7:**
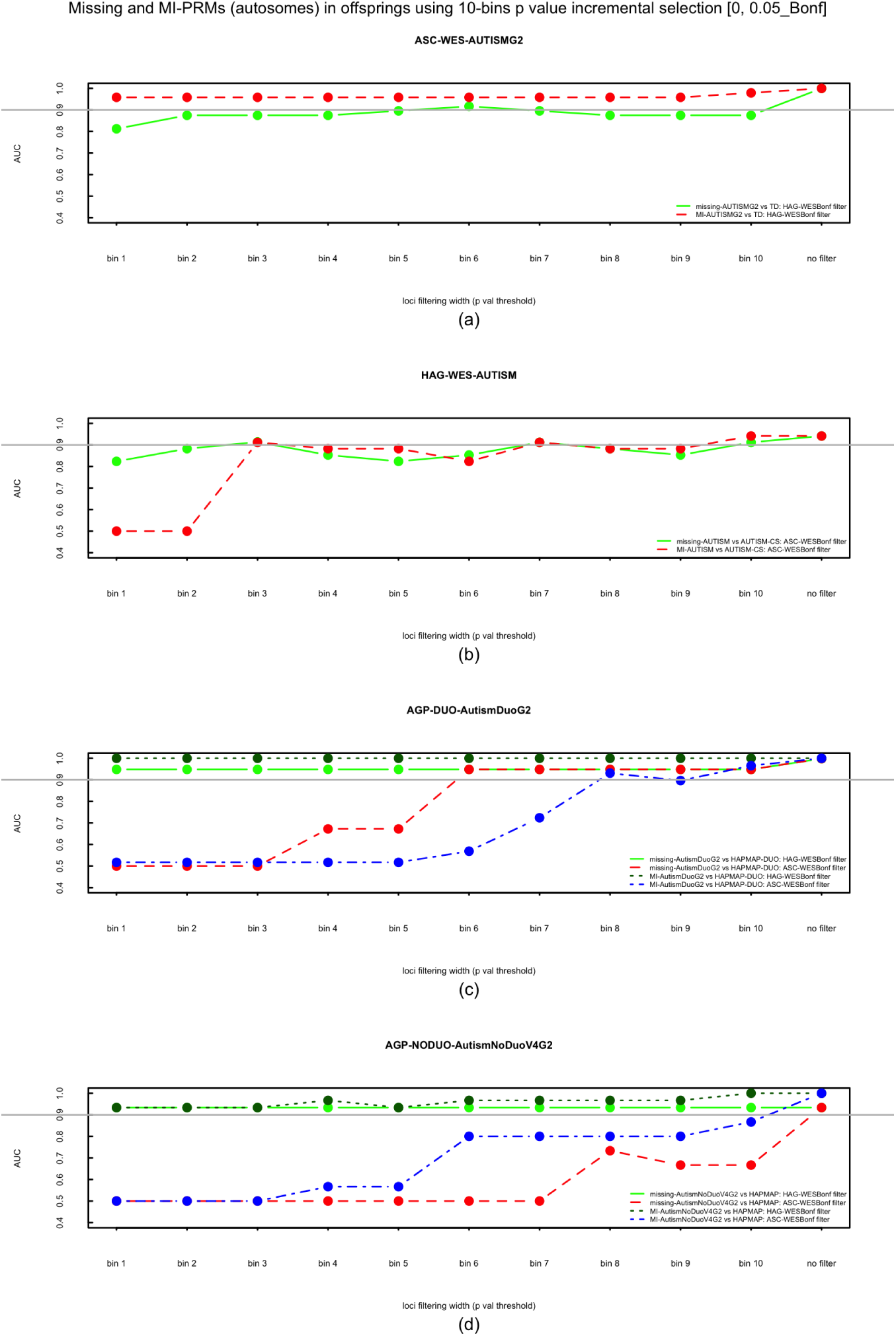
AUC using missing and MI PRMs with filtering by significant p values (*α* = 0.05, Bonf. correction) returned by the replication data subsets. Filtering was performed using AUTISM from ASC-HAG vs their control sibling subset –AUTISM-CS– (green and dark green lines for missing and MI PRMs respectively) and AUTISMG1 versus TD from ASC-WES dataset (red and blue lines for missing and MI PRMs respectively). X-axis shows results using loci bins from the lowest p-values to the highest p values (last tick is for results without filtering).

#### 1.3. Karyotypes

Either being truly associated or in linkage disequilibrium, the distribution of significant loci spreads all over the genome and many loci are significant in more than one or even all the four data sets used. We identified genes within significant loci and plot karyoplots (see figure 8 and supplementary figure S1 for missing and MI PRMs (Bonferroni correction) respectively using loci from the training/test pair subsets from ASC-WES for filtering (as it is the more dense technology used). Genes within significant loci only in ASC-WES are shown in light color (cyan for genes within loci with larger missing/MI rates in cases, pink for genes with larger missing/MI rates in controls). Genes within significant loci also in HAG-WES training/test data subsets (upper track) or training/tests subsets from arrays sources (lower track) are shown in blue (larger rates in controls) and red (larger rates in cases).

**Figure 8:**
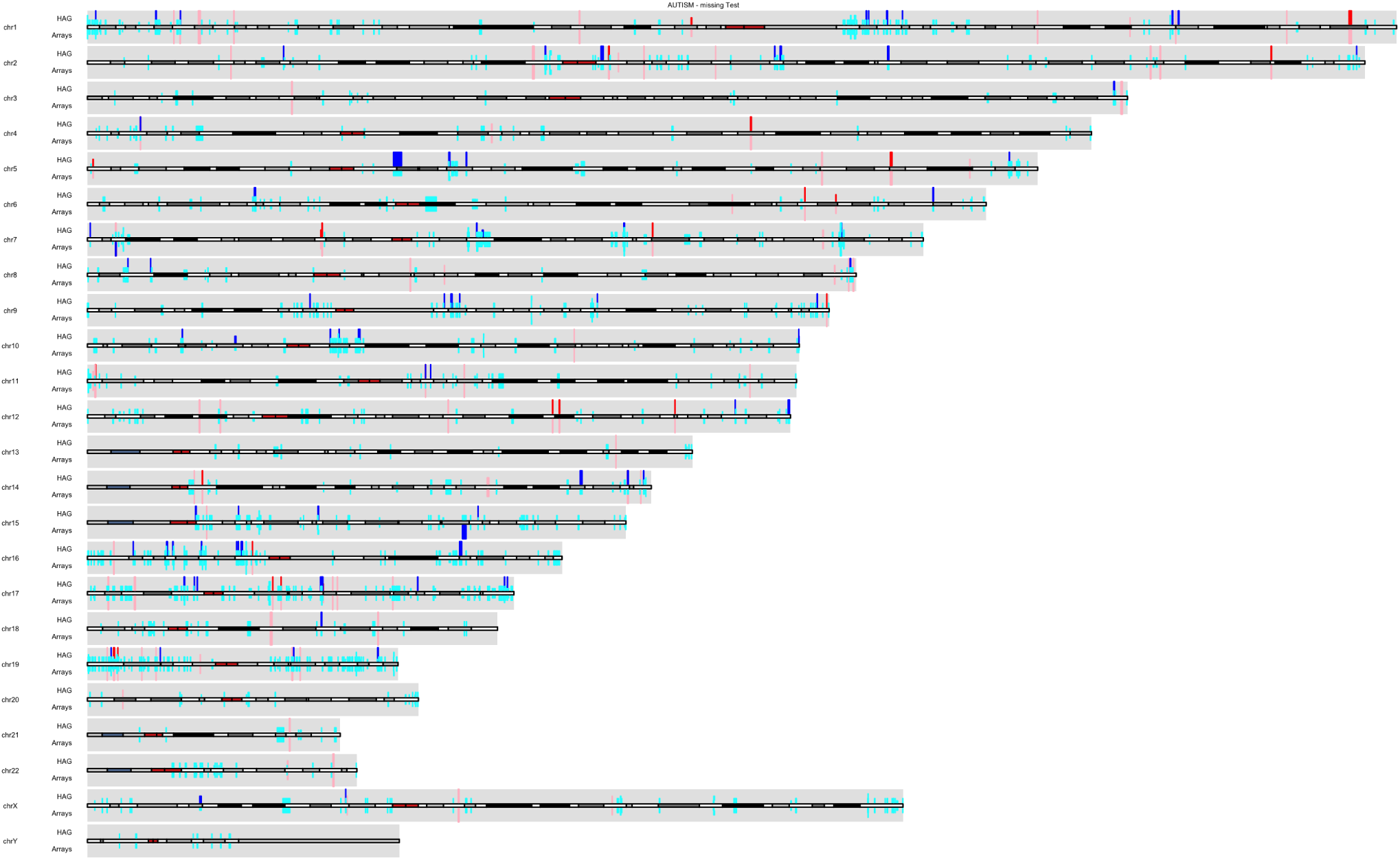
Karyoplot showing genes related to positions with *α* = 0.05 (after Bonferroni) in missing-based *χ*^2^ test for autism versus typical development offspring from AUTISMG1 vs TD (ASD-WES source) genotypes. Genes with larger proportion of missing in unaffected individuals are coloured in red. Genes with larger proportion of missing in affected individuals are coloured in blue. Genes with significant positions also in AUTISM vs AUTISM-CS (ASD-HAG source) data subsets are shown in the upper track, while genes with significant positions in any array data subsets (Human 1M and Human 1MDuo sources) are shown in the lower track (pink color if the proportion is shorter in unaffected for the dataset in a given track when was larger in the main track; cyan color if the proportion is shorter in affected for the dataset in a given track when was larger in the main track.

#### 1.4. Expression by tissues

Using the list of genes within the significant loci in autosomes from repairing-MI PRM in AUTISM versus AUTISM-CS from HAG-WES, we plotted a heatmap with the expression level in different tissues (RNA-seq data from the Human Protein Atlas Project E-MTAB-2836^17^). In this plot (figure 9) and other using alternative gene sets from other significant PRMs (data not shown) it could be observed a stronger expression level around cerebral cortex. Another clear cluster, still consistent among plots, was around testis. Similar results were obtained when using all chromosomes and only males or only females (see supplementary figures S2 and S3 respectively).

**Figure 9:**
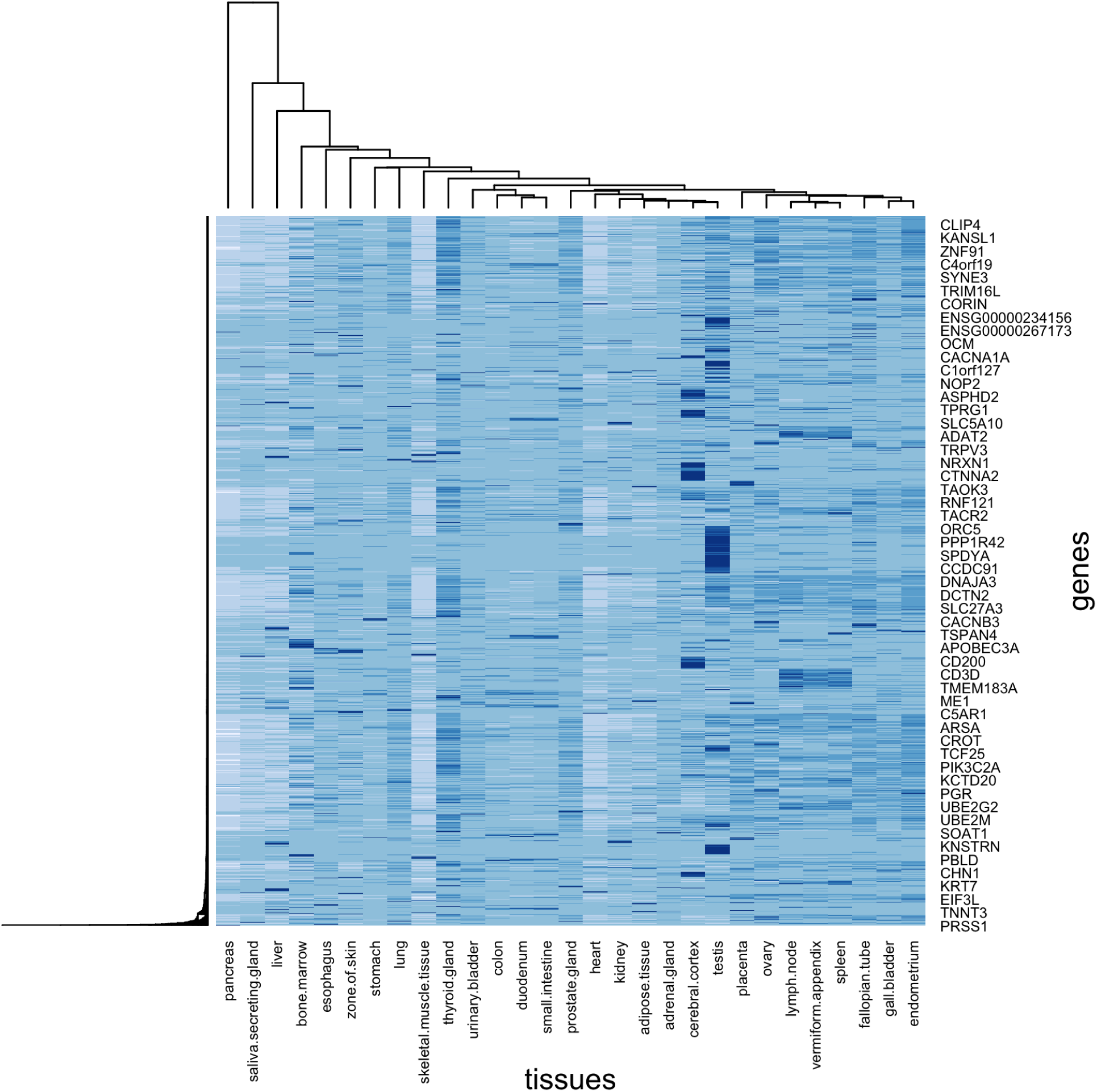
Expression heatmap using genes within significant loci for MI *χ*^2^ test (p value < 0.05, Bonf. correction) using all chromosomes (offsprings) in AUTISM vs AUTISM-CS data subsets from the HAG-WES source.

#### 1.5. Sex chromosomes

There are also a large number of statistically significant loci in sex chromosomes when comparing case/control counts required to build our different PRMs using males. As it happened in autosomes, there were agreement between datasets only within the same group of technologies (WES and genome-wide arrays). Table 6) shows results for chromosome X. We imputed individual gender by using chromosome X (to estimate inbreeding coefficients) and chromosome Y (using missing counts). We found high rates of imputation errors due to several loci in chromosome Y (missing genotypes) compatible with cnvs, or inversions, which it would point out to partial or complete sex chromosome aneuploidies in all autism data subsets in males (see table 7, which shows results for training/test and replication data subsets of autism cases within the four data sources). Results are consistent between all data subsets, having the largest rates compatible with the same haploinsufficiency karyotype: 45,X, except for those from the Human 1M Duo source (see Methods). More than 90% offspring in subsets from ASC-WES have a large amount of loci compatible with this karyotype and 84.2% in AUTISM-HAG. Results from array data (Human 1M source) are much lower (lightly above 15% in both data subsets). This significant reduction of imputation errors in autistic cases from array-based genotypes can be explained by a higher presence of duplicons or cnvs over deletions. In fact, although WES technologies would classify as missing either cnvs or deletions, the algorithm IGC used to call genotypes from array-based genomes, classifies as missing only HH genotypes, i.e., non-hybridized probes, while cnvs usually increase hybridization intensities (see methods). The conjecture of high rates of cnvs –perhaps because of duplicons–, inversions or any other structural modification instead of genetic deletions yielding to high rates of missing loci in chromosome Y (or even to consider most autistic males or *Autistic Carrier (AC)*s –fathers– without chromosome Y) is also supported by the fact that male 45,X has not been found significantly associated with autism when compared with common 46XY.^18^ Still we plot (figure 10) a comparative map of chromosome Y to observe differences in snps coverage between technologies (black segments) but, mainly, to observe differences in snps classified as missing in autistic sons (red segments) or their fathers (green segments) by ASC-WES, HAG-WES and AGP-NODUO. In agreement with results from tests used to impute sex based on chromosome Y, it can be observed by eye lower rates of missing loci in AGP-NODUO compared with WES technologies. This hypothesis of cnvs instead of deletions associated with autistic cases helps understand why there were very few significant loci in autosomes under inherited-missing PRMs in AGP-DUO and not any one in AGP-NODUO while there were thousand of them in datasets from WES technologies (see table 5).

**Figure 10:**
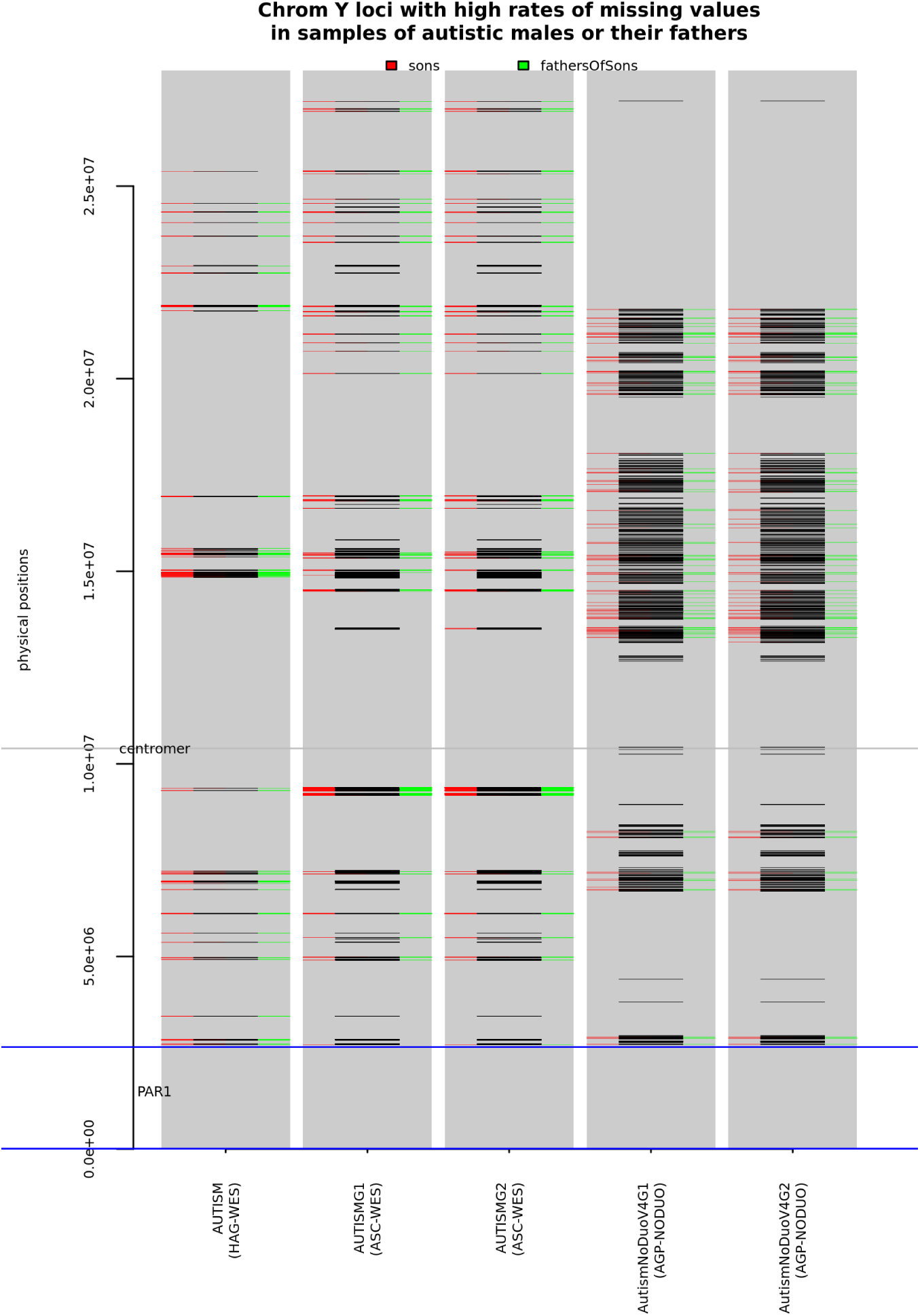
Comparative map of chromosome Y between different technologies (HAG-WES, ASC-WES and AGP-NODUO) and autistic males or their fathers in training/test (AUTISM, AUTISMG1 and AutismNoDuoG1 respectively) and replication data subsets (AUTISMG2 and AutismNoDuoG2 for ASC-WES and AGP-NODUO respectively –there is not any replication data subset for HAG-WES–) within them (X axis). Physical positions are shown in Y axis (Y range is limited to the regions genotyped by at least one of the technologies here considered). Loci coverage for each technology is shown with black segments and loci with high rates of missing genotypes are shown with red segments for cases (sons) and green segments for their fathers. PAR1 region is limited by blue horizontal lines (PAR2 is outside the map) and centromer region is marked with a grey horizontal line.

**Table 6:** Counts of significant loci within chrom X in males (p val*<* 0.05, Bonferroni correction) associated to autism (males) for the main training-test data subsets of each data source used at the training-test step for the 6 different PRMs (columns) and three different statistical tests to measure case/control differences: *χ*^2^, *χ*^2^ (Yat in the table) and N-1 two proportion (N-1 in the table) tests. Contr. (controls) means the number of significant loci under the alternative hypothesis of control (or, in general, the second data subset/trait) having higher missing/MI rates and Case (cases) means the number of significant loci under the alternative hypothesis of higher proportions of missing/MI in cases than in controls. In case of not any significant positions found at all, a hyphen is shown under both (Contr. and Case) columns. Results should be compared between tests but only within data subsets, as sample sizes are different between data subsets (see table 1) and loci counts are different between technologies (see table 2).

| Data subsets | stat. | missing |  | inhMissing |  | deNovoMissMI |  | basicMI |  | MI |  | repairingMI |  |
| --- | --- | --- | --- | --- | --- | --- | --- | --- | --- | --- | --- | --- | --- |
|  |  | Case | Contr. | Case | Contr. | Case | Contr. | Case | Contr. | Case | Contr. | Case | Contr. |
| AUTISM-HAG | $\chi^2$ | 380 | 0 | 418 | 0 | 0 | 0 | - | - | 0 | 376 | 0 | 480 |
| vs | Yat | 308 | 0 | 303 | 0 | 0 | 0 | - | - | 0 | 278 | 0 | 311 |
| AUTISM-CS | N-1 | 380 | 0 | 418 | 0 | 0 | 0 | - | - | 0 | 376 | 0 | 479 |
| AUTISMG1 | $\chi^2$ | 288 | 1 | 36 | 0 | 0 | 1 | 0 | 0 | 12 | 37 | 0 | 3 |
| vs | Yat | 206 | 1 | 14 | 0 | 0 | 0 | 0 | 0 | 1 | 17 | 0 | 2 |
| TD | N-1 | 259 | 1 | 26 | 0 | 0 | 1 | 0 | 0 | 1 | 25 | 0 | 5 |
| AutismNoDuoV4G1 | $\chi^2$ | 0 | 25 | - | - | 0 | 17 | 0 | 1 | 0 | 2 | - | - |
| vs | Yat | 0 | 3 | - | - | 0 | 3 | 0 | 0 | 0 | 2 | - | - |
| HAPMAP | N-1 | 0 | 33 | - | - | 0 | 29 | 0 | 0 | 0 | 3 | - | - |
| AutismDuoG1 | $\chi^2$ | 3 | 439 | 0 | 0 | 0 | 292 | 0 | 134 | 0 | 41 | - | - |
| vs | Yat | 2 | 80 | 0 | 0 | 0 | 47 | 0 | 110 | 0 | 25 | - | - |
| HAPMAP-DUO | N-1 | 2 | 437 | 0 | 0 | 0 | 294 | 0 | 140 | 0 | 41 | - | - |

**Table 7:** Proportions of problematic gender check in fathers and sons for cases in training/test and replication subsets –first and second rows, respectively, within each pair of consecutive rows– in the four data sources (there is not replication subset in ASC-HAG data source). Columns tXXY, tXXYY, tX and tXX mean some loci were found compatible with karyotypes 47XXY, 48XXYY, 45X and 46XX respectively.

| Dataset | father issues | tXXY father | tXXYY father | tX father | tXX father | son issues | tXXY son | tXXYY son | tX son | tXX son |
| --- | --- | --- | --- | --- | --- | --- | --- | --- | --- | --- |
| AUTISM-HAG | .819 | 0 | 0 | .807 | .012 | .842 | 0 | 0 | .842 | 0 |
| AUTISMG1 | .959 | 0 | .040 | .918 | 0 | .920 | 0 | 0 | .920 | 0 |
| AUTISMG2 | .916 | 0 | .033 | .883 | 0 | .936 | 0 | 0 | .914 | .021 |
| AutismNoDuoV4G1 | .209 | 0 | .016 | .193 | 0 | .217 | 0 | .016 | .193 | .008 |
| AutismNoDuoV4G2 | .133 | 0 | .006 | .120 | .006 | .150 | 0 | 0 | .142 | .007 |
| AutismDuoG1 | .574 | 0 | .562 | 0 | .012 | .568 | 0 | .565 | 0 | .003 |
| AutismDuoG2 | .017 | 0 | .003 | .005 | .007 | .020 | 0 | .006 | .002 | .011 |

In females, we did not found neither large rates of statistically significant loci (see supplementary table S1) or between-dataset agreement in gender imputation (see supplementary table S2), but it could be a problem of sample size, as most data sets have very few autistic females.

As both arrays used (Illumina Human 1M and Human 1M Duo) included cnvs, we built missing-based PRMs with them on sex chromosomes, including PAR, in order to observe whether there could be also significant rates of missing cnvs known from reference genomes in autistic cases. To filter loci in order to obtain those positions with the largest association with autism, we used the same approach as it was done for autosomes and snps: we considered 10 bins of loci with incremental size, with the first one with those loci with the lowest p values and the last one with all loci within p values lower than a given threshold. For generalization purposes, loci selection was performed on datasets from the other Illumina array. Figure 11 shows results from AGP-DUO (a) and AGP-NODUO (b) in males (sons).

**Figure 11:**
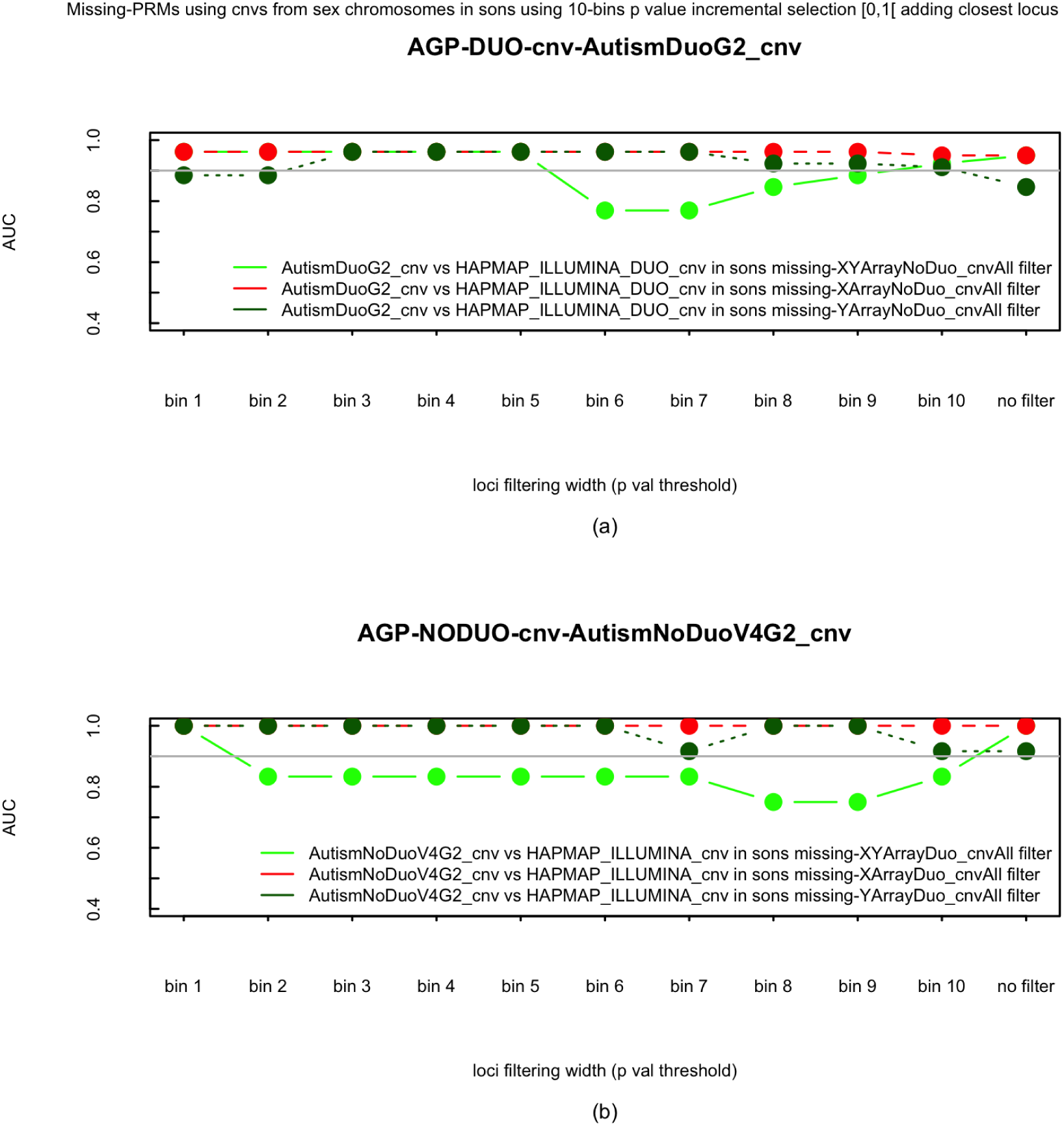
AUC using missing PRMs in cnvs datasets (Illumina arrays Human1M and Human 1M Duo, males) with filtering by significant p values (*α* = 0.05, Bonf. correction) returned by the replication data subsets. Filtering was performed using PAR (green) and non PAR (red) loci in chromosome Y from the other Illumina array. X-axis shows results using loci bins from the lowest p-values to the highest p values (last tick is for results without filtering). AUC are obtained from the replication data subsets. (a) Results from Human 1M Duo array (AGP-DUO) using cnv genotypes from AutismDuoG1 (cases) and HAPMAP-ILLUMINA-DUO (controls) for training/test and AutismDuoG2 (cases) as replication data subset. Loci selection was obtained using data from AutismNoDuoV4G1 and HAPMAP-ILLUMINA. (b) Results from Human 1M array (AGP-NODUO) using cnv genotypes from AutismNoDuoV4G1 (cases) and HAPMAP-ILLUMINA (controls) for training/test and AutismNoDuoV4G2 (cases) as replication data subset. Loci selection was obtained using data from AutismDuoG1 and HAPMAP-ILLUMINA-DUO.

Figure 12 shows results from AGP-DUO (a) and AGP-NODUO (b) in females (daughters). AGP-NODUO reaches maximum AUC from the first bin in the three filters (chromosome X, chromosome Y and PAR) in sons (*AUC* = 1 in the three filters) and daughters (*AUC* = 0.777 in the three filters). AGP-DUO reaches the maximum AUC at first bin with chromosome X and PAR filters and at third bin with chromosome Y filter, having in the three cases the same value *AUC* = 0.9615 in sons. In daughters, *AUC* = 0.9166 in X and PAR filters and *AUC* = 1 in Y filter.

**Figure 12:**
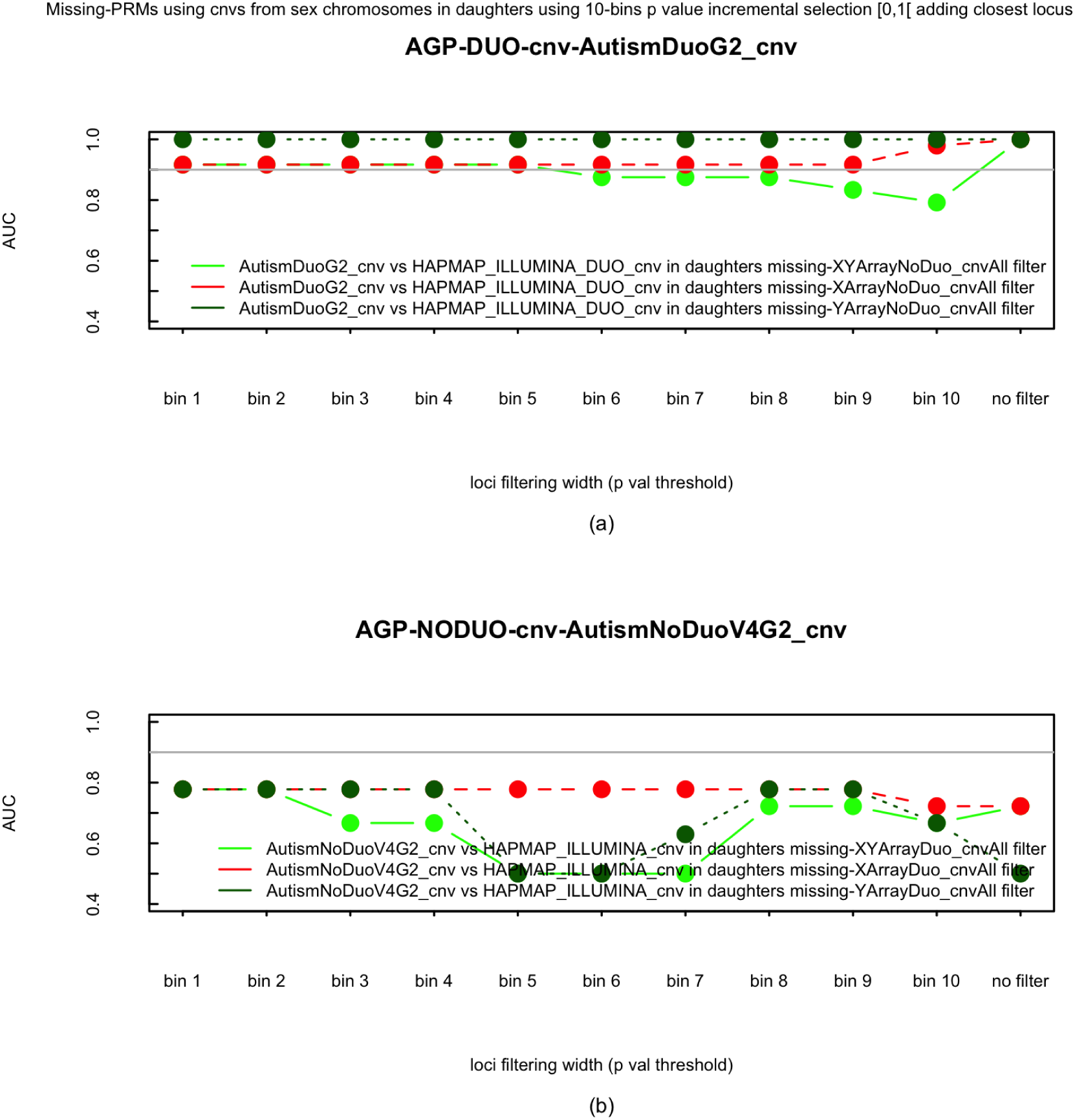
AUC using missing PRMs in cnvs datasets (Illumina arrays Human1M and Human 1M Duo, females) with filtering by significant p values (*α* = 0.05, Bonf. correction) returned by the replication data subsets. Filtering was performed using PAR (green) and non PAR (red) loci in chromosome Y from the other Illumina array. X-axis shows results using loci bins from the lowest p-values to the highest p values (last tick is for results without filtering). AUC are obtained from the replication data subsets. (a) Results from Human 1M Duo array (AGP-DUO) using cnv genotypes from AutismDuoG1 (cases) and HAPMAP-ILLUMINA-DUO (controls) for training/test and AutismDuoG2 (cases) as replication data subset. Loci selection was obtained using data from AutismNoDuoV4G1 and HAPMAP-ILLUMINA. (b) Results from Human 1M array (AGP-NODUO) using cnv genotypes from AutismNoDuoV4G1 (cases) and HAPMAP-ILLUMINA (controls) for training/test and AutismNoDuoV4G2 (cases) as replication data subset. Loci selection was obtained using data from AutismDuoG1 and HAPMAP-ILLUMINA-DUO.

When looking for loci agreement in the selected genes between arrays, we found, in sons, agreement in PAR: both arrays reach maximum *AUC* = 1 with only one and the same locus: rs17884601 –GRCh37:1449813– belonging to gene IL3RA (interleukin 3 receptor subunit alpha) in PAR1. We also found agreement in chromosome Y: maximum *AUC* = 0.9615 in Human 1M Duo reached at bin 3 with 2 loci selected –rs13304367 and rs7893075–, all of them within gene PRKY (pseudogene protein kinase Y-linked); and maximum *AUC* = 1 in Human 1M reached at bin 1 with only one locus selected, rs13304367, within gene PRKY as well. In chromosome X, maximum *AUC* = 1 is reached at bin 1 in both arrays, but there is not any locus in common. In daughters we found agreement in PAR as well, although not as good as with sons: maximum AUC is reached at bin 1 in both arrays, being *AUC* = 0.9166 in Human 1M Duo and *AUC* = 0.7777 in Human 1M, with one locus in common, rs17884601 –the only one in Human 1M Duo–, belonging to gene IL3RA in PAR1, and a second locus in Human 1M, rs17884601, belonging to gene IL9R (interleukin 9 receptor) in PAR2. There was also agreement in chromosome X: same AUCs as in PAR, reached at bin 1 as well, with 5 loci –rs17264623, rs7472606, rs35327279 –within gene PCDH11X (protocadherin 11 X-linked)–, rs35133165 –within gene PCDH11X as well– and rs5983864 –within gene CLIC2 (chloride intracellular channel 2)– selected by Human 1M Duo and 49 in Human 1M, with all 5 loci selected by Human 1M Duo also included in the 49 ones selected by Human 1M. In chromosome Y, maximum AUC is reached at bin 1 in both arrays: *AUC* = 1 with 23 loci selected by Human 1M Duo and *AUC* = 0.7777 with 22 loci selected by Human 1M, but there were few genes in common.

In summary, a cnv (rs17884601) within gene IL3RA allows perfect classification of affected and unaffected sons, in both datasets, according to whether individuals have not any allele at all (autistic male offspring from Human 1M and Human 1M Duo arrays) or have a variable number of alleles (pseudo-control male offspring from HAPMAP CEU with the same arrays). It has to be noted (figure fig:replicationChromYsons) that AUC decreases right within the next bin. Gene IL3RA is known to be expressed in human brain development by influencing human brain volume variation.^19^ In fact, macrocephaly is associated with autism.^20^

Cnvs within gene PRKY allows almost perfect, but not perfect, classification. This, together with the fact that PRKY is very close to PAR1, may make this gene not to be within the origin of autism but, because of its strong linkage disequilibrium with IL3RA, help increase phenotypic diversity and additive effect of autism. In fact, abnormal recombination with its homologous (PRKX, within chromosome X) is known to be the cause of one third of sex aneuplodies causing sex chromosome reversal (XX males, XY females) .^21^ This may explain why we found so many autistic individuals with (at least multiple short-range chromosome-wide) sex aneuploidies and why, mainly in Human 1M Duo, complete (long-range) XX males and XY females were directly observed from gentprints (an XX male is the father within the trio gentprint in figure S4).

In females, autism cannot be explained only by deletions within gene IL3RA. Even with deletions within gene IL9R in PAR2, AUC is still very low in Human 1M array. These results may also explain why autism is more common in males than in females, as females also require deletions of genes within PAR2, and, as these genes are located in PAR allowing genetic interchange between gender, why autism affects both genders.

## Discussion

We believe that results here presented open a different approach to build genetic risk scores to predict individual risk of autism and other complex diseases in general. By using missing and MI patterns we have confirmed was it was known about high heritability and disease prevalence rates in autism. Without considering sex chromosomes, and when using incremental loci selection according to p values or the outperforming autosome, AUC is above 0.9 in replication datasets with affected and unaffected individuals, including a dataset with unaffected siblings, which probes the genetic origin of autism. There are no differences between affected offspring and their parents (AUC 0.5), a fact that confirms autism as a complex disease, i.e., a trait that requires not only to carry the appropriate genetic variants but some environmental factor for the disease onset. By using a cnv missing-based risk model, we have identified a unique gene, IL3RA in PAR1, able to explain the whole variance (*AUC* = 1) in males, instead of the large amount of snps required to reach the maximum AUC in autosomes. We also observed chromosome-wide short-range aneuplodies in sex chromosomes (see table 7), detected more frequently in WES than in array-based datasets, explaining missing loci in cases as rare duplications or cnvs instead of deletions. These, at least short-range aneuplodies, may be explained because another gene, PRKY, very close to PAR1 and therefore in strong linkage disequilibrium with IL3RA, is associated to sex aneuplodies and was found highly associated with autism in males (deletion of a cnv in this gene gave AUCs larger than 0.9 in both arrays). In females, it was also necessary but not sufficient a cnv deletion within gene IL9R in PAR2 to explain the whole variance. AUC results from autosomes (see figure 7) were also very high but they usually required more loci to reach its maximum value within each series, which may point out to several loci in linkage with sex chromosomes, and therefore, contributing to autism. This is also supported by the fact that associated genes in autosomes are mostly expressed in cerebral cortex and testis (see figure 9). Therefore, we have also confirmed the polygenetic etiology of autism, with hundred thousand variants having an additive effect in autism, or rather, its polygenetic variability because of strong linkage in sex chromosomes and autosomes with the true cause in gene IL3RA. This also could explain the high differences in autism prevalence by gender. In fact, the origin in the deletion of a cnv in IL3RA in males is transmitted to females because of PAR1 recombination, but females require more genetic variants, including the gene IL9R in PAR2, which belongs to the second, more recent, PAR.

In light of these results, the explanation of DNVs as possible genetic causes ^22,23^ or any other genetic cause not detected by a missing or MI based PRM, still would require to have inherited an important amount of rare variants from parents, only present at such given rate in ACs.

While there are no differences between affected individuals and their parents (AUC *≈* 0.5), affected individuals can be clearly differentiated from their unaffected full siblings (AUC above 0.9). How DNA repair can be done all over the chromosome in healthy individuals (we found more than 10% of loci where DNA repair could have occurred in healthy siblings, according to positive test under the repairing MI PRMs in HAG data subsets) is an open question. The existence of long-range linkage disequilibrium between autosomes and sex chromosomes would not explain healthy siblings from AC. One hypothesis is that short-term duplicons or cnvs, which may extend along all autosomes,^24^ were linked to sex chromosomes during meiosis, perhaps due to translocations, and transmitted to AC offspring.

There is an ethical question that we need to consider, regarding practical applications: the development of a risk predictor of autism, using either genome arrays or genome sequencing, may become soon. It will be very important to understand, at least in light of these results, that Asperger are genetically closer to TD than to autism (perhaps is because of this that Asperger is usually considered not a disease but a peculiar trait). The same may be true for other ASD sub-groups, such as PDD-NOS (see figure 5). The development of tests able to distinguish between strict autism and other ASDs will become very important, specially because people with these peculiar traits may have a highly satisfactory and independent life.^25^

## Data Availability

All data results produced in the present work are contained in the manuscript.
The software we developed is available online at
https://bios.ugr.es/IGC/

## Data availability

The datasets used for the analysis described in this manuscript were obtained from the IHPM^14^ and from db-GaP at http://www.ncbi.nlm.nih.gov/gap through dbGaP accession number, phs000267.v5.p2 (AGP consortium) and phs000298.v4.p3 (ASC).

## Code availability

The code of software IGC for array genotyping and all the code used for data preparation and analysis are available upon request by contacting MariaMar Abad-Grau at.

## Acknowledgments

Array-based datasets used in this work were obtained from dbGaP at http://www.ncbi.nlm.nih.gov/gap through dbGaP accession number, phs000267.v1.p1. Submission of the data, phs000267.v1.p1, to dbGaP was provided by Dr. Bernie Devlin on behalf of the Autism Genome Project (AGP). Collection and submission of the data to dbGaP were supported by a grant from the Medical Research Council (G0601030) and the Wellcome Trust (075491/Z/04), Anthony P. Monaco, P.I., University of Oxford.^26,27,28^

ASC-WES datasets used in this work were obtained from dbGaP at http://www.ncbi.nlm.nih.gov/gap through db-GaP accession number, phs000298.v4.p3. This work was supported by National Institutes of Health (NIH) grants U01MH100233, U01MH100209, U01MH100229 and U01MH100239 to the Autism Sequencing Consortium. Sequencing at Broad Institute was supported by NIH grants R01MH089208 (M.J.D.) and sequencing by U54 HG003067 (S. Gabriel, E. Lander) and UM1HG008895 (Gabriel, Lander). We acknowledge the clinicians and organizations that contributed to samples used in this study. Finally, we are grateful to the many families whose participation made this study possible.

HAG-WES datasets used in this work were obtained from dbGaP at http://www.ncbi.nlm.nih.gov/gap through dbGaP accession number, phs000639.v3.p1. Sequencing data from this study was generated with support from grants from the National Institute of Mental Health (MH083565; MH089952) and the Simons Foundation to Christopher A. Walsh at Boston Children’s Hospital.

## 2. Supplementary Material

**Figure S1:**
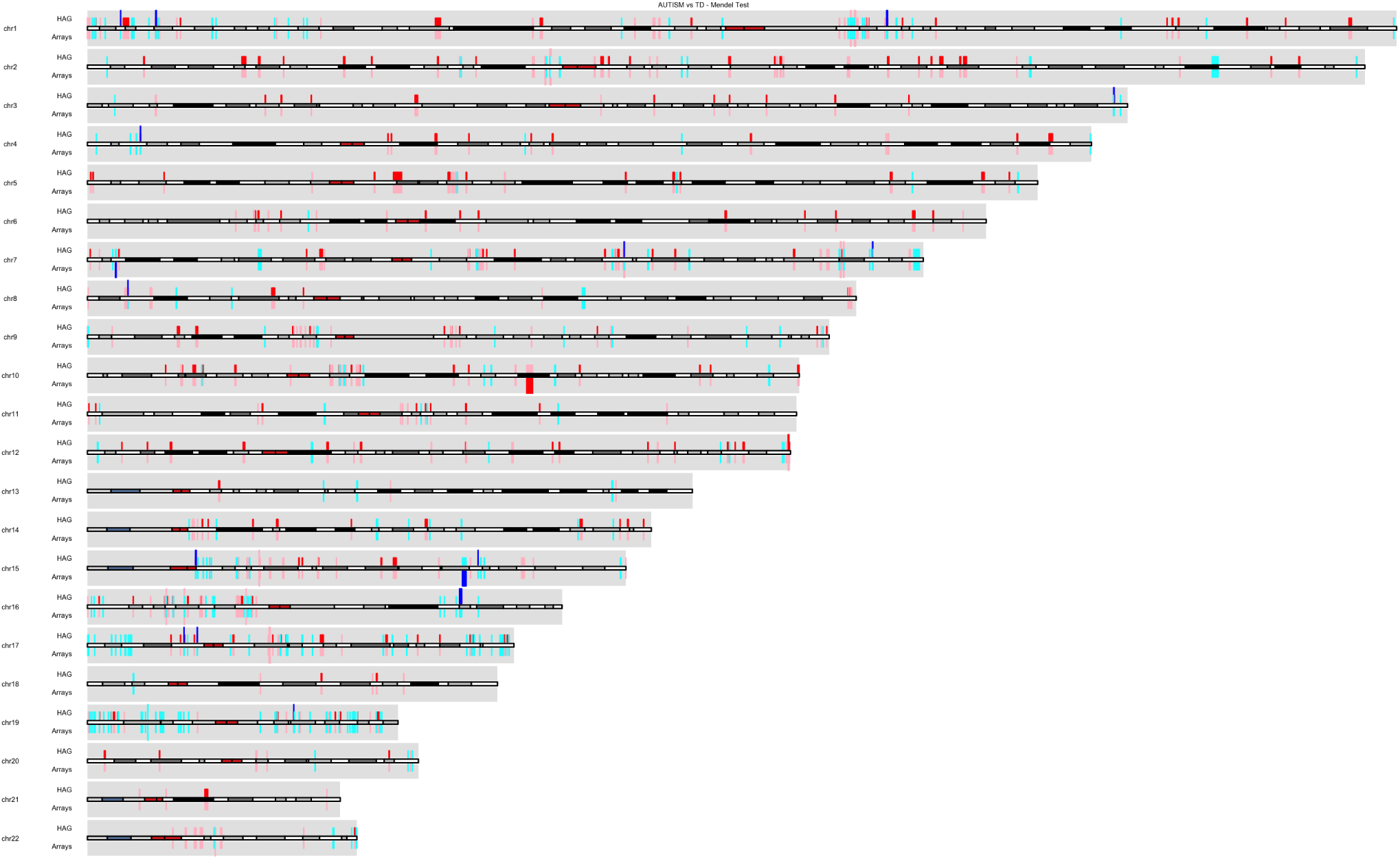
Karyoplot showing genes related to positions with *α* = 0.05 (after Bonferroni) in MI-based *χ*^2^ test for autism versus typical development offspring from AUTISMG1 vs TD (ASD-WES source) genotypes. Genes with larger proportion of MI in unaffected individuals are coloured in red. Genes with larger proportion of MI in affected individuals are coloured in blue. Genes with significant positions also in AUTISM vs AUTISM-CS (ASD-HAG source) data subsets are shown in the upper track, while genes with significant positions in any array data subsets (Human 1M and Human 1MDuo sources) are shown in the lower track (pink color if the proportion is shorter in unaffected for the dataset in a given track when was larger in the main track; cyan color if the proportion is shorter in affected for the dataset in a given track when was larger in the main track.

**Figure S2:**
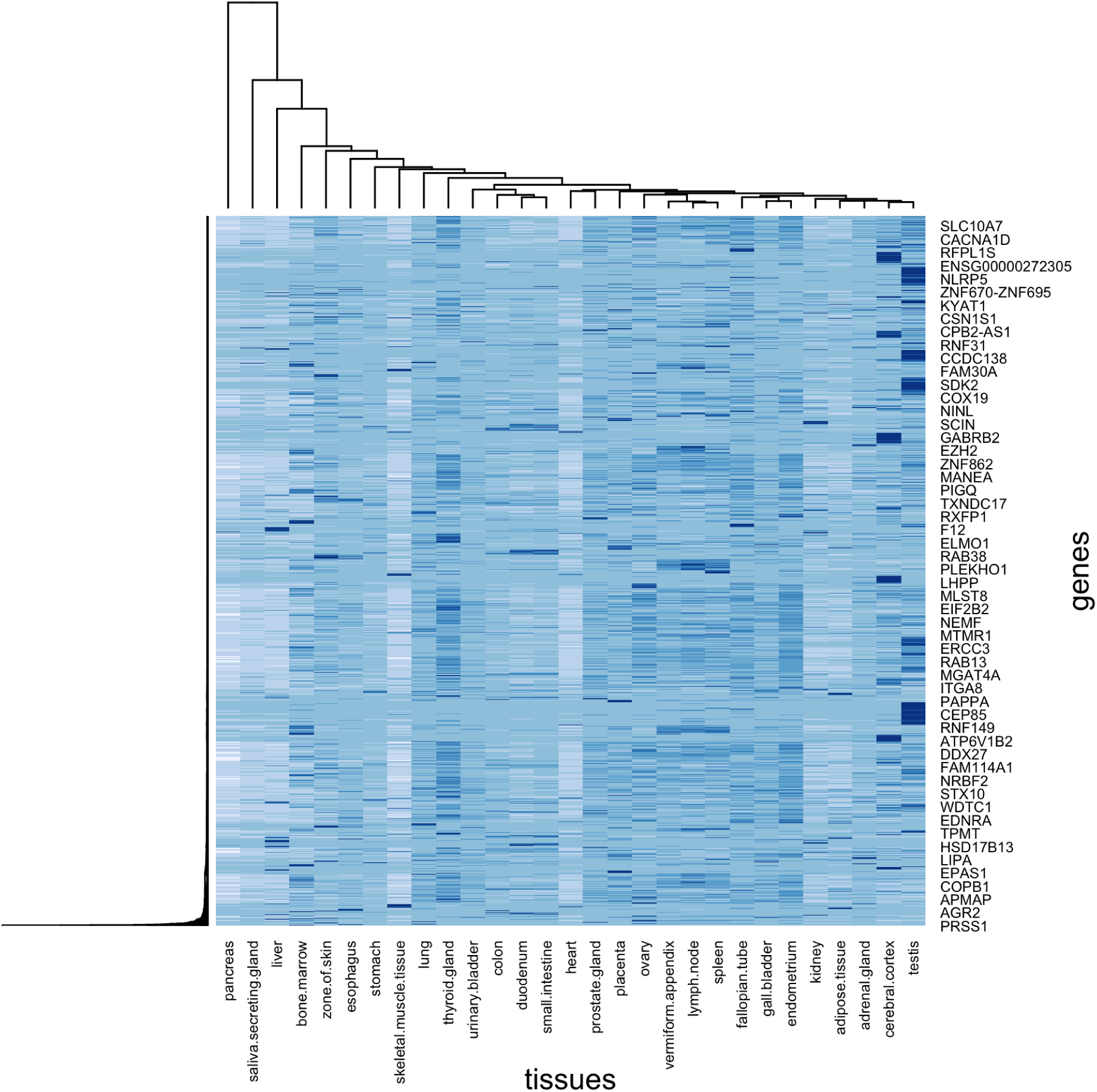
Expression heatmap using genes within significant loci for MI *χ*^2^ test (p value < 0.05, Bonf. correction) using all chromosomes (sons) in AUTISM vs AUTISM-CS data subsets from the HAG-WES source.

**Figure S3:**
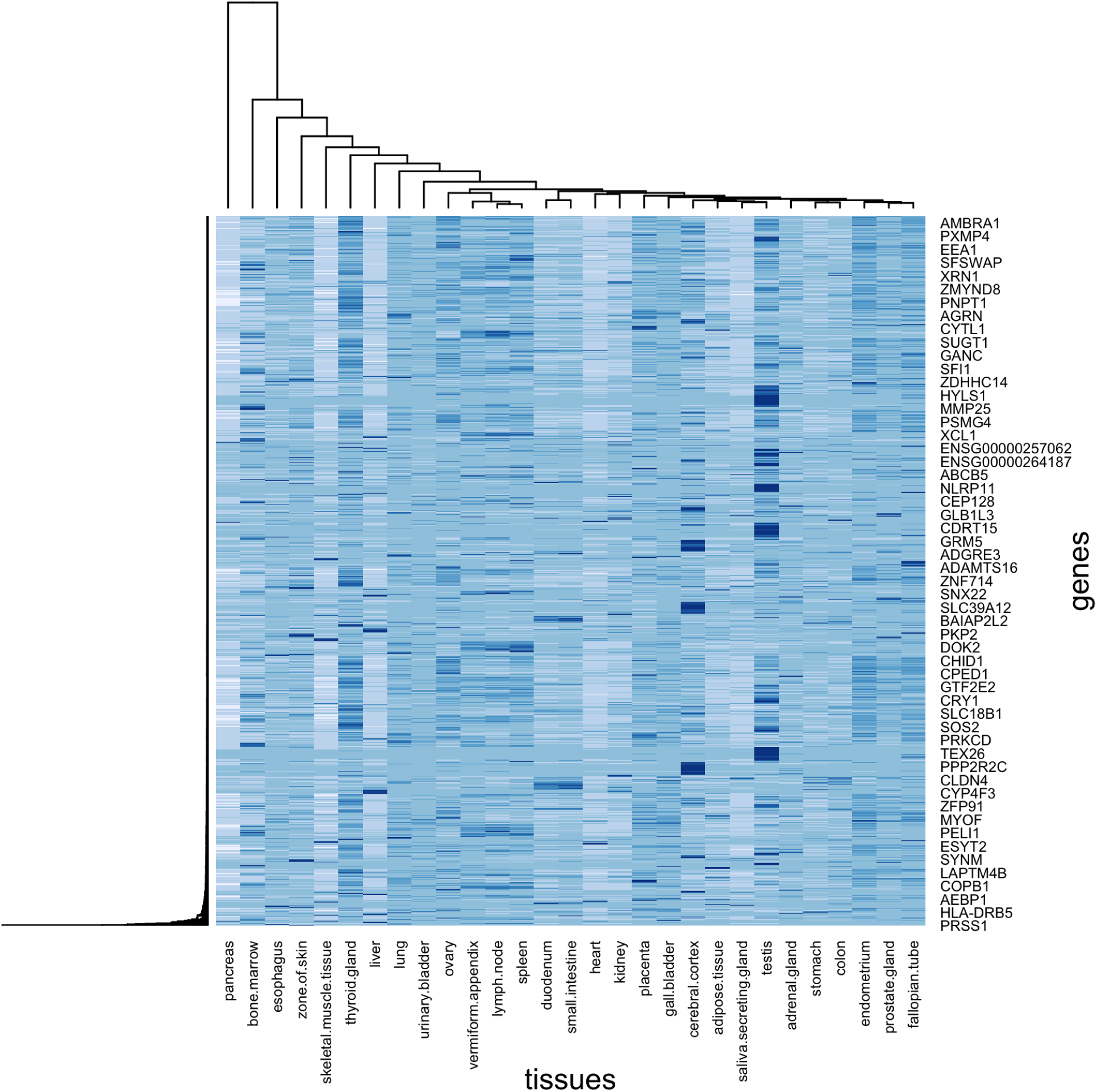
Expression heatmap using genes within significant loci for MI *χ*^2^ test (p value < 0.05, Bonf. correction) using all chromosomes (daughters) in AUTISM vs AUTISM-CS data subsets from the HAG-WES source.

**Figure S4:**
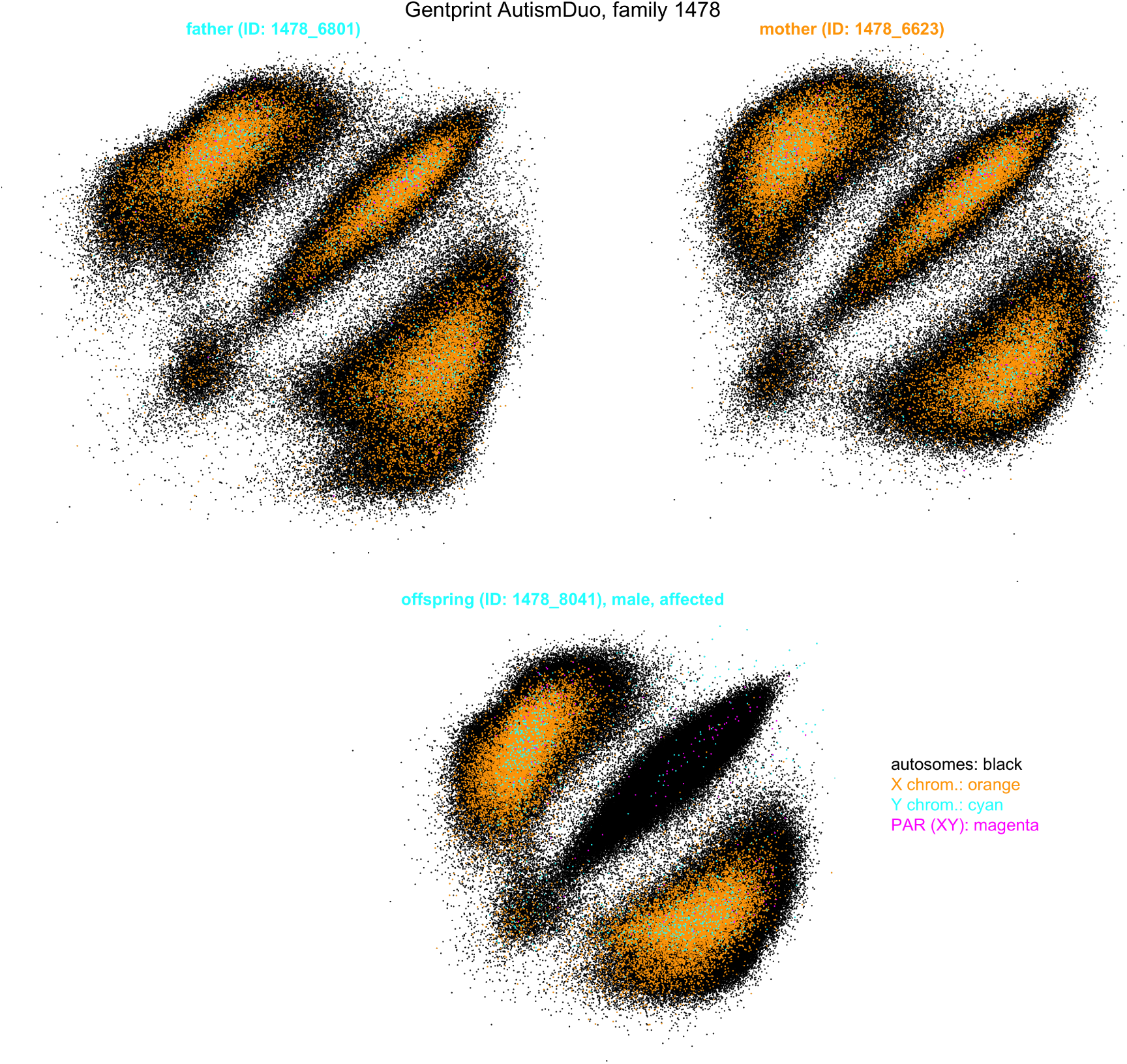
Gentprints (log of raw allele intensities) from a male (at the bottom) and her parents (father on the left, mother on the right). Axis range are the same for the three family members. X chrom intensities are plot in orange and Y chrom intensities are plot in cyan. There are a large amount of heterozygous Y chromosome (cyan) at least in both parents, what it could be compatible with the Klinefelter syndrome. Father also shows a female pattern for X chromosome (XX chromosome), wile mother shows a male pattern for Y chromosome or even YY genotypes. Therefore, both parents seem to have XXYY genotypes. Offspring, male, seems to be more compatible with the expected XY genotype. There are also very large amounts of unknown genotypes in the three members of the family (what we have called HH cluster). The father –and more lightly the child– also shows what seems to be compatible with chromosome-wide copy number variations (very long haplotypes repeated) as it can be observed like two overlapping AA and BB clusters (homozygotic genotypes).

**Figure S5:**
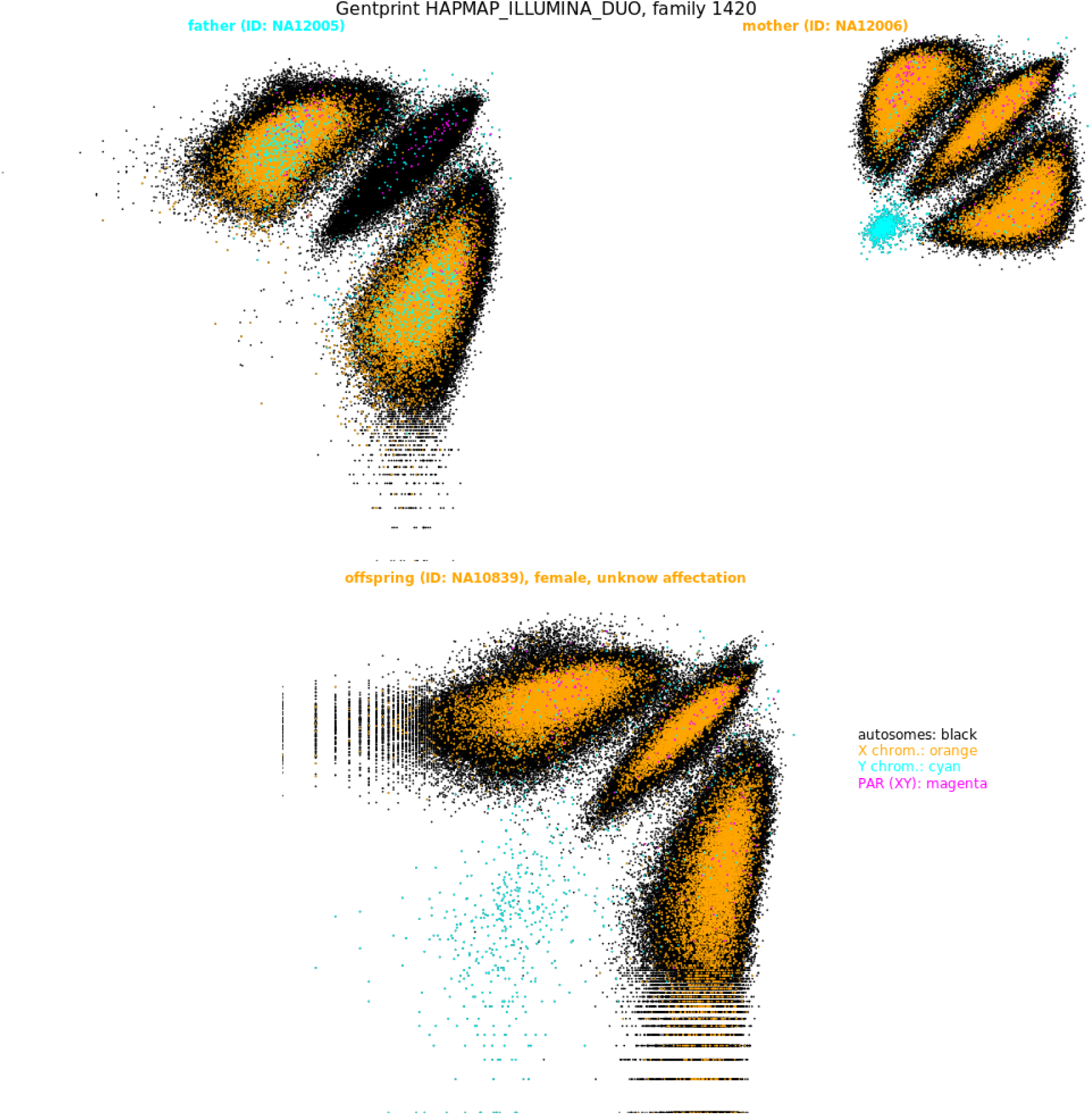
Gentprints (log of raw allele intensities) from a daughter (at the bottom) and her parents (father on the left, mother on the right). Axis range are the same for the three family members. It can be observed large differences among them, not only in intensity ranges but in shapes.

**Table S1:**
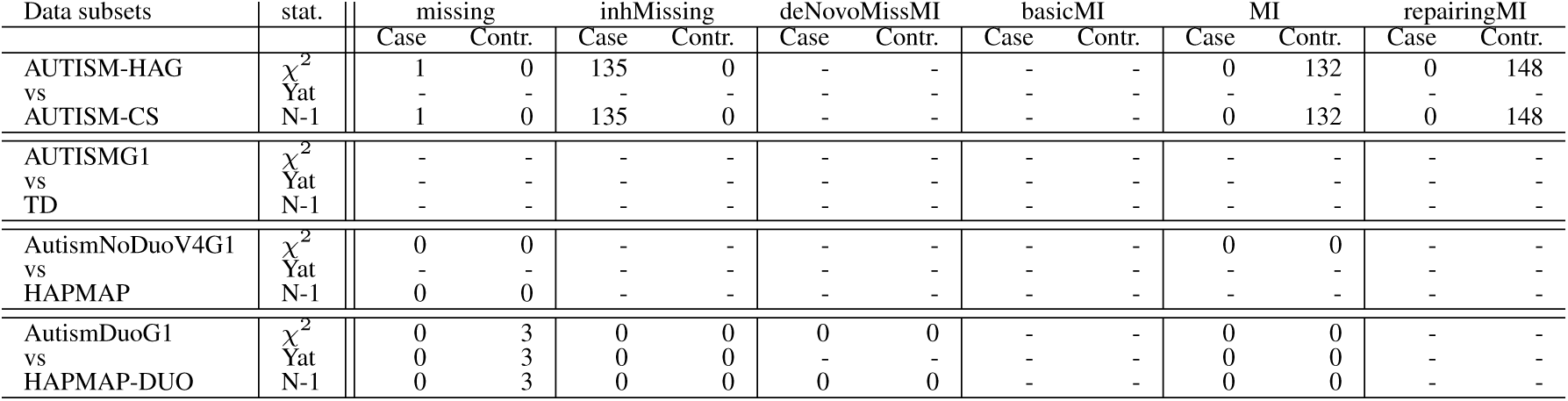
Counts of significant loci within chrom X in females (p val*<* 0.05, Bonferroni correction) associated to autism (females) for the main training-test data subsets of each data source used at the training-test step for the 6 different PRMs (columns) and three different statistical tests to measure case/control differences: χ^2^, 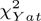 (Yat in the table) and N-1 two proportion (N-1 in the table) tests. Contr. (controls) means the number of significant loci under the alternative hypothesis of control (or, in general, the second data subset/trait) having higher missing/MI rates and Case (cases) means the number of significant loci under the alternative hypothesis of higher proportions of missing/MI in cases than in controls. In case of not any significant positions found at all, a hyphen is shown under both (Contr. and Case) columns. Results should be compared between tests but only within data subsets, as sample sizes are different between data subsets (see table 1) and loci counts are different between technologies (see table 2).

| Data subsets | stat. | missing |  | inhMissing |  | deNovoMissMI |  | basicMI |  | MI |  | repairingMI |  |
| --- | --- | --- | --- | --- | --- | --- | --- | --- | --- | --- | --- | --- | --- |
|  |  | Case | Contr. | Case | Contr. | Case | Contr. | Case | Contr. | Case | Contr. | Case | Contr. |
| AUTISM-HAG | $\chi^2$ | 1 | 0 | 135 | 0 | - | - | - | - | 0 | 132 | 0 | 148 |
| vs | Yat | - | - | - | - | - | - | - | - | - | - | - | - |
| AUTISM-CS | N-1 | 1 | 0 | 135 | 0 | - | - | - | - | 0 | 132 | 0 | 148 |
| AUTISMG1 | $\chi^2$ | - | - | - | - | - | - | - | - | - | - | - | - |
| vs | Yat | - | - | - | - | - | - | - | - | - | - | - | - |
| TD | N-1 | - | - | - | - | - | - | - | - | - | - | - | - |
| AutismNoDuoV4G1 | $\chi^2$ | 0 | 0 | - | - | - | - | - | - | 0 | 0 | - | - |
| vs | Yat | - | - | - | - | - | - | - | - | - | - | - | - |
| HAPMAP | N-1 | 0 | 0 | - | - | - | - | - | - | - | - | - | - |
| AutismDuoG1 | $\chi^2$ | 0 | 3 | 0 | 0 | 0 | 0 | - | - | 0 | 0 | - | - |
| vs | Yat | 0 | 3 | 0 | 0 | - | - | - | - | 0 | 0 | - | - |
| HAPMAP-DUO | N-1 | 0 | 3 | 0 | 0 | 0 | 0 | - | - | 0 | 0 | - | - |

**Table S2:** Proportions of problematic gender check in mothers and daughters for cases in training/test and replication subsets –first and second rows, respectively, within each pair of consecutive rows– in the four data sources (there is not replication subset in ASC-HAG data source). Columns tXXY, tX and tXY mean some loci were found compatible with karyotypes 47XXY, 48XXYY, 45X and 46XY respectively.

| Dataset | mother issues | tXXY mother | tXXYY mother | tX mother | tXY mother | daughter issues | tXXY daughter | tXXYY daughter | tX daughter | tXY daughter |
| --- | --- | --- | --- | --- | --- | --- | --- | --- | --- | --- |
| AUTISM-HAG | 0 | 0 | 0 | 0 | 0 | .041 | 0 | 0 | .041 | 0 |
| AUTISMG1 | .083 | 0 | 0 | 0 | .083 | 0 | 0 | 0 | 0 | 0 |
| AUTISMG2 | 0 | 0 | 0 | 0 | 0 | .071 | 0 | 0 | 0 | .071 |
| AutismNoDuoV4G1 | .038 | 0 | .038 | 0 | 0 | .038 | 0 | .038 | 0 | 0 |
| AutismNoDuoV4G2 | .040 | 0 | .040 | 0 | 0 | 0 | 0 | 0 | 0 | 0 |
| AutismDuoG1 | 1.000 | 0 | 0 | 0 | .620 | 1.000 | 0 | 0 | 0 | .689 |
| AutismDuoG2 | .041 | 0 | .023 | 0 | .017 | .035 | 0 | 0 | 0 | .035 |

